# Visualising candidate behaviour in computer-based testing: Using *ClickMaps* for exploring *ClickStreams* in undergraduate and postgraduate medical examinations

**DOI:** 10.1101/2023.06.13.23291148

**Authors:** I. C. McManus, Liliana Chis, Albert Ferro, S. Helen Oram, James Galloway, Vikki O’Neill, Gil Myers, Alison Sturrock

**Author notes:** Educational advisor, MRPC(UK). Head of Research Unit, MRCP(UK). Associate Director for Written Examinations, MRCP(UK).

## Abstract

**Background:** The rapid introduction of computer-based testing (CBT) in UK (United Kingdom) undergraduate and postgraduate medical education, mainly as a result of the COVID-19 pandemic, has generated large amounts of examination data, which we call the *ClickStream*. As candidates navigate through exams, read questions, view images, choose answers and sometimes change answers and return, re-read, and make further changes, the multiple actions are recorded as a series of time-stamped clicks or keystrokes. Analysing that mass of data is far from simple, and here we describe the creation of *ClickMaps*, which allow examiners, educationalists and candidates to visualise behaviour in examinations.

**Methods:** As an example of *ClickMaps*, we describe data from a single examination lasting three hours, with 100 ‘best-of-five’ questions, which was one of two papers sat in 2021 by 508 candidates as a part of the MRCP(UK) Part 2 exam. Two *ClickMaps* were generated for each candidate. The *Full ClickMap* allows the complete three-hours of the examination to be visualised, while the *Early ClickMap*, shows in more detail how candidates responded during the first six minutes of presentation of each of the 100 questions in the exam.

**Results:** Since the primary purpose of this paper is expository, detailed descriptions and examples of *ClickMaps* from eleven candidates were chosen to illustrate different patterns of responding, both common and rare, and to show how straightforward are *ClickMaps* to read and interpret.

**Conclusions:** The richness of the data in *ClickStreams* allows a wide range of practical and theoretical questions to be asked about how candidates behave in CBTs, which are considered in detail. *ClickMaps* may also provide a useful method for providing detailed feedback to candidates who have taken CBTs, not only of their own behaviour but also for comparison with different strategies used by other candidates, and the possible benefits and problems of different approaches. In research terms, educationalists urgently need to understand how differences in *ClickMaps* relate to differences in student characteristics and overall educational performance.

## Background

Although computer-based testing (CBT; also computer-delivered assessment, CBA) for knowledge-based assessments in medicine had gradually become more popular over the past two decades [1], with many advantages over pencil-and-paper assessments [1], with high acceptance by candidates, although less so by educators [2]. CBT in the UK developed suddenly and extensively as a result of the COVID-19 pandemic. Repeated lockdowns and a need for internet-based remote examinations accelerated the use of CBT, and with that growth came an abundance of rich data on the second-by-second behaviour of examination candidates as they answered questions in exams that lasted several hours. That mass of information, which we call the *ClickStream*, shows there to be wide variations in how candidates behave in exams, an area which previously could not easily be studied. However, making sense of raw *ClickStream* data is a challenge for examiners and educators, and here we describe *ClickMaps*, visual summaries of candidate behaviour in CBT exams which are straightforward to interpret and may provide useful feedback to candidates on how their behaviour differs from that of other candidates.

CBT exams can take many forms, from formats which are essentially reproductions of pencil-and-paper exams with multiple-choice questions (MCQs), and other formats such as very short answers (VSAs) [3, 4]. CBTs can also vary in their invigilation styles, some using remote proctoring, while other are open-book assessments. *ClickMaps* can in principle be applied to all of them, where the task is to understand how candidates make decisions in what usually is a time-limited environment.

Computers have been used for marking multiple-choice questions in medical examinations since the 1960s, albeit that initially the answers had to be transferred by hand onto 80-column punch cards [5], but subsequently optically scanned answer sheets were developed [6].

Such exams, which were still ‘pencil-and-paper’ tests from a candidate’s perspective, were used in medicine for most knowledge assessments for the next three or four decades. True CBT, where candidates interact directly with computers when answering questions, was developed in the 1990s, partly due to cheaper computers and developing internet connectivity. The United States Medical Licensing examination (USMLE) went online in 1999 [7], and some undergraduate examinations also explored the use of CBT [8]. Nevertheless a 2004 review in the BMJ still described CBT as “an emerging technology with great potential”, but acknowledged that, “implementation can be challenging and lengthy”, and, “the costs and expertise necessary … should not be underestimated” [7]. The potential for statistical analysis of detailed timing measures from CBTs was shown by the 2001 detailed modelling of response times in USMLE, finding effects of item ordering, difficulty, length and complexity of wording, as well as candidate knowledge of English [9].

The present group of authors, who work on undergraduate and postgraduate assessments in several different UK medical schools, had separately begun interpreting the complex data from CBTs, and in discussion jointly began developing and exploring the concept of *ClickMaps*. Some of that work had begun pre-COVID, as for instance with keystroke data from MRCP(UK) Speciality Certificate Examinations, which have used CBT since their inception in 2009, but keystroke data were mostly used for studying anomalous behaviour and collusion, as well as for analyses of question quality [10]. A current challenge is that there are many online exam providers using different software with different candidate interfaces, options and displays, which export data in different formats with different computer codes describing candidate behaviour. As an introduction for examiners, educationalists and candidates, we here describe examples of *ClickMaps*, for simplicity using examples from a single examination, the MRCP(UK) Part 2 examination, which we hope show the potential of *ClickMaps* for exploring and understanding candidates’ exam behaviour in CBT assessments. In this paper the term ‘click’ refers generically to both mouse clicks, keyboard responses and navigation actions using the cursor, as well as other codes generated within the examination software.

## Methods

The MRCP(UK) diploma has three parts, Part 1 and Part 2 which are both knowledge assessments [11], and PACES which is an assessment of clinical skills [12, 13]. The Part 2 examination currently consists of two papers, each containing 100 questions and lasting three hours, using single-best-answer questions (SBAs; Best of Five, BoF), but results from only one of the two papers is shown here. The CBT version of the examination, which was implemented in 2020 during the first UK pandemic lockdown to replace the previous pencil-and-paper assessment, was sat remotely using a candidate’s own computer. The software ensured that no other programmes, such as for accessing the internet, could be accessed while taking the examination. Invigilation was by remote proctoring, the proctor being able to see the candidate via their computer’s video camera. Registration and timing for the exam was carried out by the software, with identity checks carried out by proctors. All examinations were video-recorded. Some candidates started the exam late because of technical issues and therefore some exams last a little longer than the standard three hours. Some candidates with reasonable adjustments or additional learning needs were allowed longer than the standard three hours, but they were not included in the diet described. An open question for CBT in general, although not considered here, is the extent to which extra time is used by candidates, the differences in which they use that time, and the potential benefits.

Examination *items* are given a *canonical (standard) numbering*, with the numbering identical for all candidates, and labelled here as C1 to C100, although in practice these may be arbitrary codes such as AN/15494 used in the question bank. The canonical item numbers are the same for all candidates, but are not displayed to candidates. The *actual presentation order* of the items, as displayed, was randomised for each candidate, so that, say, canonical item C5 could be presented anywhere from the first question (Q1) to the last, 100^th^, question (Q100), so that each item was equally likely to appear as a question in any particular presentation order. *Items* here refers to canonical items and *questions* to items in the order as presented. For this exam, the five possible answers for questions are presented in the same order for all candidates, typically alphabetical (although some exams do choose also to randomise answer orders across candidates).

All items were presented as a stem (which for Part 2 typically was quite lengthy), followed by a question, usually fairly brief, and the five possible answers. Of the 100 items, many included numerical data, such as biochemistry results, and 20 (20%) included images, such as ECGs, X-rays, etc, which appeared in a separate window when candidates clicked to see them, with codes in the *ClickStream* indicating when the image window was being viewed.

The exam software provided navigation buttons so candidates could select answers, move on to new questions, ‘flag’ or ‘pin’ questions to be returned to, and go back or forward to answers which were flagged, which had been left unanswered, or for a general review of previous answers. A clock display of remaining time was also provided on the screen. An example test, and other information on exam formatting etc, is available at https://www.mrcpuk.org/mrcpuk-examinations/part-2/uk-online-exam.

*ClickMaps* were generated using software written in *R* [14], which used the graphics package *ggplot2* [15]. For present purposes, *ClickMaps* were created for the 508 candidates who took Paper 1 of a Part 2 exam sat in 2021. The *ClickMaps* shown here have been anonymised so that individual candidates cannot be identified, and information on overall performance is presented in terms of approximate percentiles, to prevent reidentification. Raw *ClickMaps* are generated in PNG format at very high resolution (18,000 x 24,000 pixels) so that examiners can zoom in on details as required.

For the present paper the tops of displays have been cropped to remove candidate information. Individual *ClickMaps* (both full and early) are typically about 5Mb in size, so that an exam with 500 candidates and two separate papers generates about five gigabytes of images. The additional files for this paper contain the original but anonymised high-resolution *Full ClickMaps* and *Early ClickMaps* for each of the eleven candidates A to K which are also shown in figures 1 to 14.

**Figure 1:**
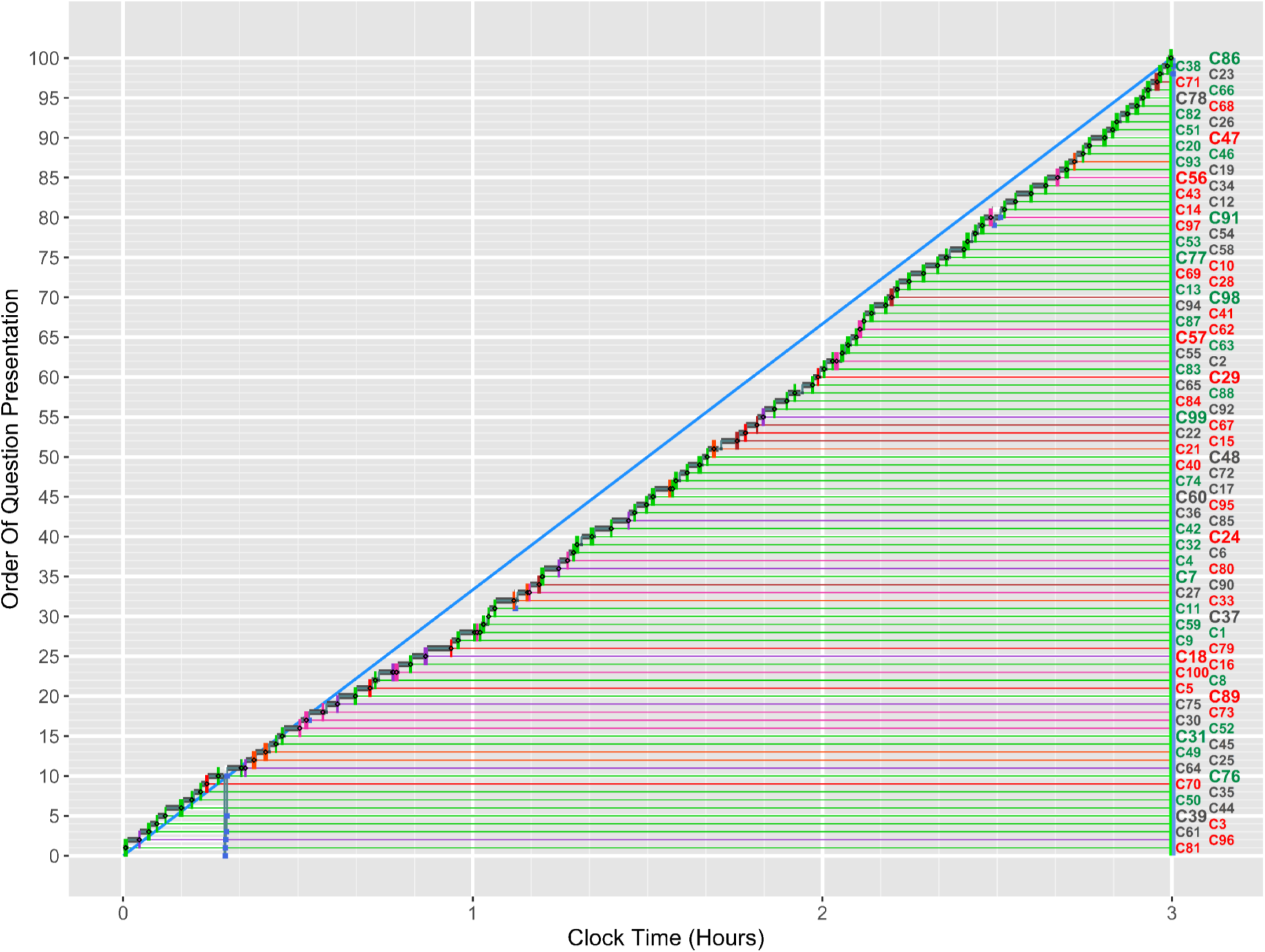
Example of a *Full ClickMap* for candidate A. See text for further details.

### Participants

Participants were candidates taking a diet of the MRCP(UK) Part 2 examination in 2021.

### Statistical analysis

Statistical analysis consisted entirely of descriptive methods using graphical techniques.

## Results

The raw *ClickStream* data analysed here are selected from 508 candidates with a total of 629,269 records, each record being an event or an action during the exam, either by a candidate or by the system. There was a mean of 1,238 records per candidate (median=1,187, SD=248, range=761 to 2,385), an average question having about 12 recorded events, which can include navigation codes, answer choices, opening images, and pinning/unpinning items. Candidates in this particular diet, during the Covid pandemic, were restricted to those who did not require additional time beyond the standard three hours. *ClickMaps* were generated for each candidate, and the entire set examined visually. Some patterns were more frequent than others, and the examples shown here were chosen both as being more convenient for explanation and being relatively simple to read and explain. Examples are not representative of the proportions of *ClickMap* types in the dataset overall, being chosen primarily for expository purposes.

### The Full ClickMap

Figure 1 shows, for Candidate A, an example of a ***Full ClickMap*** which displays the entire three hours of the exam and shows a pattern of responding which is fairly common amongst candidates. For Candidate A, and for all the candidates shown in the figures the high resolution versions of the *Full* and *Early ClickMaps* are available in the additional files (see Additional Files 1 and 2, and files 3 to 22 for other candidates).

Figure 2 shows an enlarged portion of the *ClickMap* in Figure 1, which can be zoomed into because of the high resolution of the raw *ClickMap*. The horizontal axis shows clock time within the exam (i.e. 0 hours is when the exam started), with thick vertical white lines indicating hours, and thin vertical white lines at ten minute intervals. The vertical axis shows the presentation order for the questions, 1 being the first question with which the candidate is presented, through to 100 which is the last question as presented. The numbers on the right-hand side indicate the canonical numbers for the questions, this candidate seeing C81 first, C96 second, C61 third, etc, through to C86 as the last question. Comparison of this *ClickMap* with later ones for other candidates shows that the questions are in a different random order for each candidate (e.g. for Candidate B, in figure 4, the 1^st^, 2^nd^, 3^rd^ and last questions presented are C53, C95, C4 and C14).

**Figure 2:**
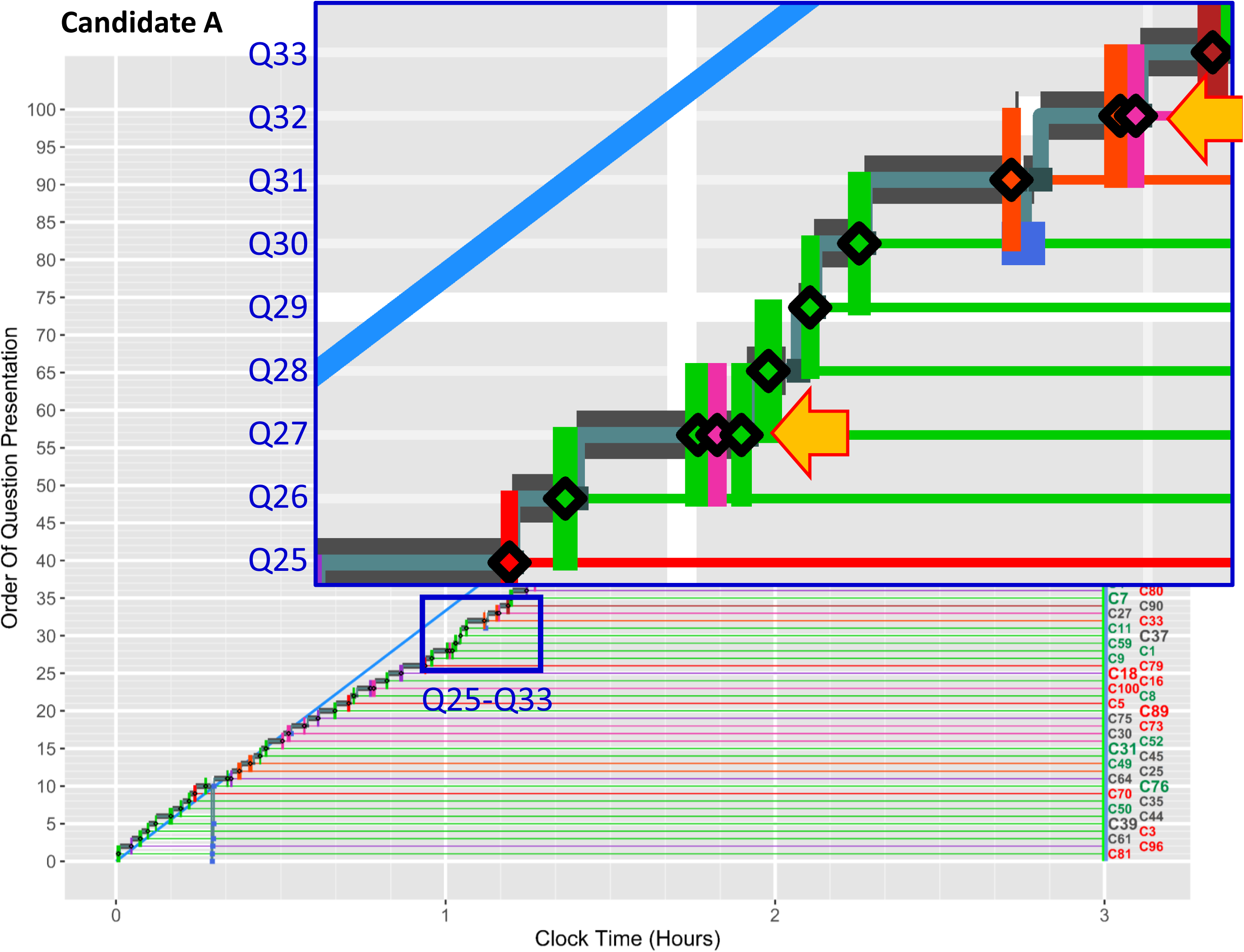
*Full ClickMap* for candidate A showing detail of Q25 to Q33. See text for further details.

The *ClickMap* shows the time at which a candidate moves to a new question, with a vertical coloured line indicating that an answer has been given, green for a correct answer and one of four reddish shades (red, orange, maroon and pink) for each of the four possible incorrect answers. For a particular canonical item, red, orange, maroon and pink might indicate, say, wrong answers A, B, D and E (with C being correct and in green); that allows comparison of wrong answer choices across candidates, and it can also show when a candidate changes a response from one incorrect answer to a different incorrect answer, or perhaps changes their response to a wrong answer that had been selected previously.

Once an item is answered then a green or reddish line extends horizontally across the *ClickMap*, making right and wrong answers easier to see. Candidate A answered 68 items correctly, as shown by the predominance of green in the diagram. A’s performance is above average, at about the 65^th^ percentile.

The line, from bottom left to top right, joining together the sequential responses of the candidate to the questions shows the progression through the questions. We call this line the **main sequence**, after the Hertzsprung-Russell diagrams used in astronomy [16]. Without defining a main sequence further, it typically represents the time during which a candidate has sequentially visited all (or perhaps most) of the 100 questions in the exam in their presentation order (but see later for more complex examples).

A challenge for candidates in any timed assessment is to ensure that all questions are answered within the set time limit. For a three-hour exam with 100 questions there is an average time of 108 seconds for answering each question, which is shown in figure 1 by the slope of the diagonal blue line from bottom left to top right. For the first half hour the main sequence of candidate A follows the blue line. The slope of the main sequence then decreases as it drops below the blue diagonal line for the next hour and a half, indicating that questions are being answered too slowly to complete them all within the time allowed. In the final hour of the exam the candidate accelerates, indicated by the increasing slope of the main sequence, and all 100 questions are just finished within the three hours.

The *ClickMap* for Candidate A, in Figure 1, provides a ‘big picture’ of the way in which a candidate approaches the exam. The resolution of the image file is however high (see additional file 1), which makes it possible to look in detail at parts of the *ClickMap*, an example being shown in figure 2. Questions 25-33, indicated in the lower blue box, are magnified within the larger blue box. The point at which an item is answered is shown by the black diamonds, with coloured bars extending above and below the question to make responses more visually obvious when viewing the *Full ClickMap*. Solid grey lines at the beginning of a question indicate that the candidate has moved to that item but has not yet made an answer. Items such as Q29 are answered quickly and correctly, whereas items such as Q26 take slightly longer, and items such as Q31 are looked at for a long while, before being answered wrongly. Most questions are answered only once, but Q 27 and Q32, indicated by orange arrows, are answered several times. Q27 is answered correctly (green line), then changed to an incorrect answer (pink line), and then back to the correct answer (green line). Q32 is answered twice, first with a wrong answer (red line) and then with a different wrong answer (pink line). These are both examples of what we call **early changes**, occurring during the first viewing of a question. *ClickMaps* for further candidates will also show examples of **late changes**.

### Early ClickMaps

Figure 2 suggests that much is occurring in the first few minutes of looking at a question, and that is better seen in what we call ***Early ClickMaps***, as in figure 3, which show the first six minutes of looking at each question. The *Early ClickMap* in figure 3 is for the same candidate as in figures 1 and 2. The six-minute window is adequate for most candidates, as the average available time for a question is 108 seconds, indicated by the vertical blue line in figure 3, meaning that six minutes is more than three times the average available time per question. The thick vertical white lines are at intervals of 30 seconds, with thin vertical white lines at 15 second intervals.

**Figure 3:**
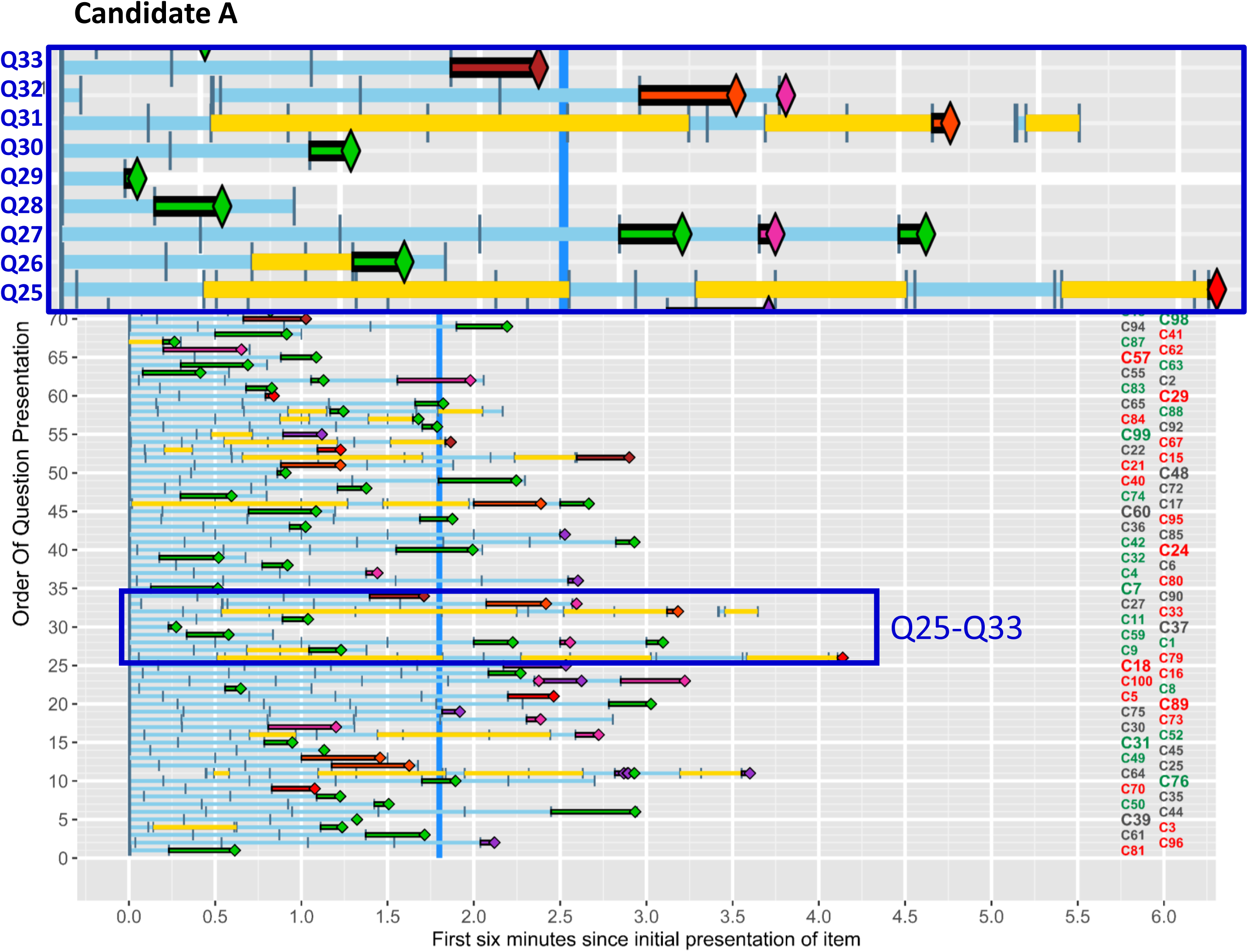
*Early ClickMap* for candidate A. See text for further details.

Conceptually, figure 3 can be seen as equivalent to figure 2, with all the questions on the vertical axis in the order of presentation in both figures, but in the *Early ClickMap* the bars for each question have been slid to the left so that times are relative to zero, the time that the item was first seen (rather than indicating exam clock time as in the *Full ClickMap*). The horizontal time axis has also been enlarged so that the first six minutes are more easily visible. The *Early ClickMap* shows more clearly the detailed internal structure of the response to each item, rather than the global, overall pattern of responses that is shown in the *Full ClickMap*. Each row in the *Early ClickMap* can be viewed as a magnification of the equivalent row in the *Full ClickMap*.

To help the present explanation, questions 25-33 in figure 3, indicated by the blue box in the lower part of the figure, are magnified at the top of the diagram, and superimposed on what normally would be questions 71 to 100. The layout of the *Early ClickMap* is somewhat different from that of the *Full ClickMap*, the main difference being that distance from the left-hand edge, indicated on the the x axis, shows time after a question is *first* looked at, rather than time after the exam started. Other differences are shown in Q25, which Candidate A looked at for over four minutes. The blue bars indicate that the candidate is looking at the text of the question, while the yellow bars indicate looking at a separate on-screen window which typically includes an X-ray or other visual material. For Q25, candidate A reads the text for 30 secs, examines the image for 75 seconds, then reads text for 20 seconds, looks at the image for 45 seconds, reads the text for a third time for about 35 seconds, and then for a third time looks at the image, for 30 seconds; and only after that do they then choose the wrong answer, shown in red. Not all image questions are as difficult, and Q26 is answered correctly in about 75 seconds, with only about 20 seconds spent looking at the image. Q31, also an image question, shows the image being looked at for a total of about 140 secs, and the text for only about 45 seconds. Interestingly, about 15 seconds after answering this question, the candidate returns to the question and its image, looks at it for nearly 15 seconds, and then does not make any further change, perhaps having checked a particular detail or interpretation.

The *Early ClickMap* also clarifies what Candidate A did in figure 2 with Q27 and Q32, for which there were three and two answers respectively were chosen. In figure 3, Q27 was first answered correctly after about 135 seconds, then, after a further 30 seconds a wrong answer was chosen, and about 35 seconds after that the candidate returned to the original answer which was correct. Not all intervals between changed answers are as long, and for Q32 there was only about ten seconds between the first (wrong) answer and the second (different wrong) answer.

### Question difficulty

Exam questions differ in difficulty. For the 100 questions in the current paper, the percentage of candidates answering questions correctly varied from 6.1% (the hardest question) to 90.4% (the easiest question), with a mean of 65.2% correct (median= 66.1%, SD=21.3%). Difficulty of items in the *ClickMaps* is shown by the colour of the canonical question numbers on the right-hand side, green indicating the easiest third of questions, grey the middle third of questions, and red the hardest third of questions. Note that red and green here do not indicate that the question has been answered correctly or incorrectly.

### Revisiting questions

The *Full ClickMap* in figure 1 shows a pattern that is typical of many candidates, most or all questions being visited only once. Other patterns though are found.

A problem for candidates using the answering pattern of figure 1 is that the candidate leaves no time to revisit questions, and perhaps to revise answers. Candidate B, in figure 4, is a relatively weak candidate, performing below the 10^th^ percentile. The main sequence shows the candidate speeding up after about one and a half hours, thereby creating some time to return to questions, the main sequence finishing in about 160 minutes, and so twenty minutes remained for returning to questions, which is about 13 seconds per question. A **secondary sequence** can be seen running through all 100 questions in the final twenty minutes of the exam, with some being looked at for longer than others. Four items had answers changed on a later viewing of the question, which we call **late changes**, and are indicated by orange arrows. These late changes are sometimes occurring more than two hours after giving the original answer. Four answers were changed, one from right to wrong, one from wrong to right (shown in the inset at top left for Q16), and two answers changed from one wrong answer to a different wrong answer.

**Figure 4:**
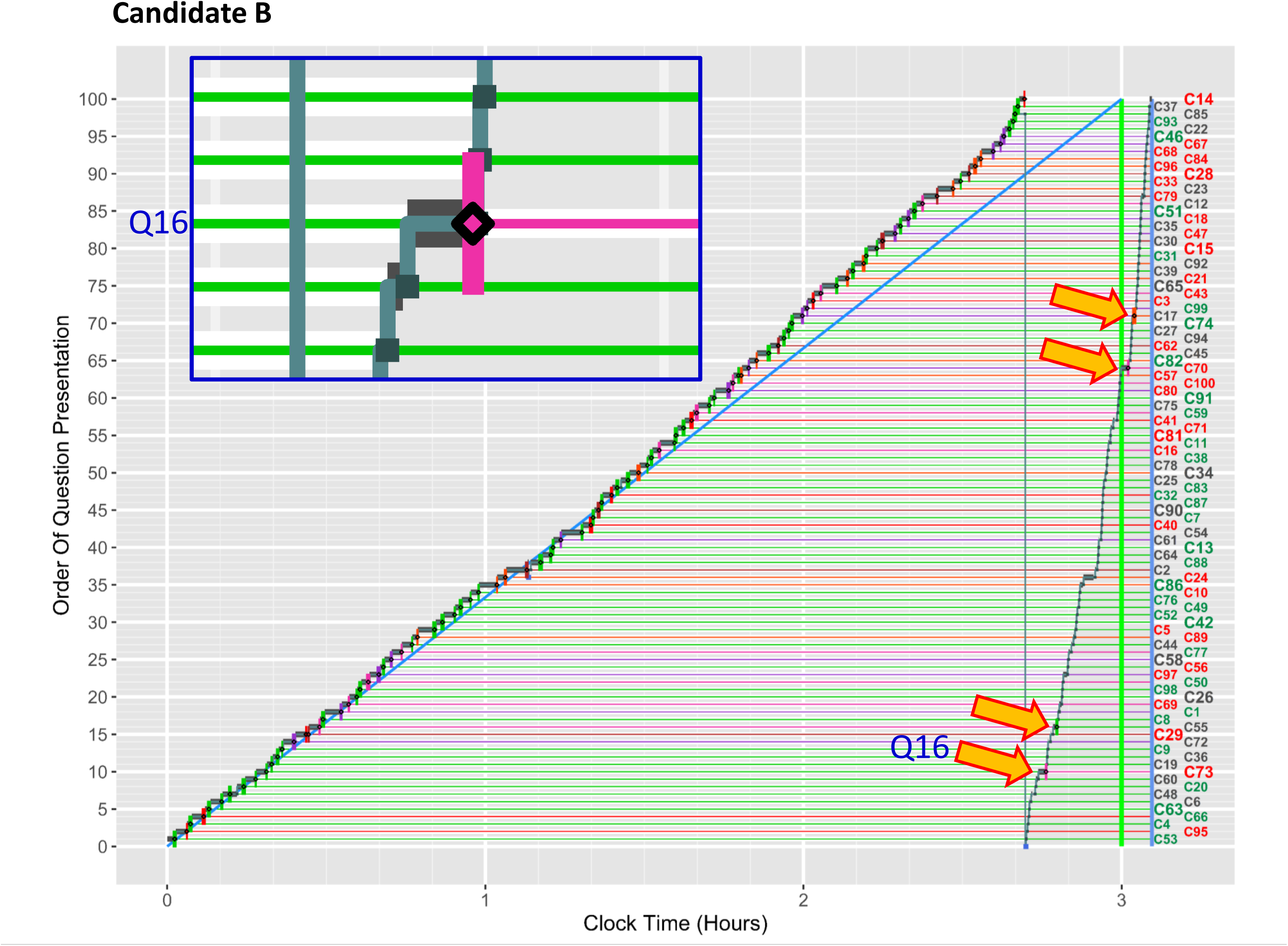
*Full ClickMap* for Candidate B including enlarged detail of response to Q16. See text for further details.

### Leaving the exam early

Although working quickly through an exam can leave time for checking previously answered questions, as with candidate B, not all candidates avail themselves of the opportunity to return to questions, as shown by Candidate C in figure 5. The main sequence is very fast, all 100 questions being answered within about 2 hours 10 minutes, leaving 50 minutes free, about 30 seconds per question, which could have been used to revisit items. However, after the main sequence finishes, candidate C returns very briefly to question 19, makes a double late change, from a wrong answer to the right answer and then changes back to the original wrong answer, at which point the candidate leaves the exam. The regulations for online examinations allow candidates to leave the exam whenever they wish, and a number choose to leave early. Candidate C was an above average candidate, on about the 70^th^ percentile, as shown by the number of green lines. Whether they would have gained a higher mark had they returned to further items is unclear, but the answer is probably yes (see Discussion).

**Figure 5:**
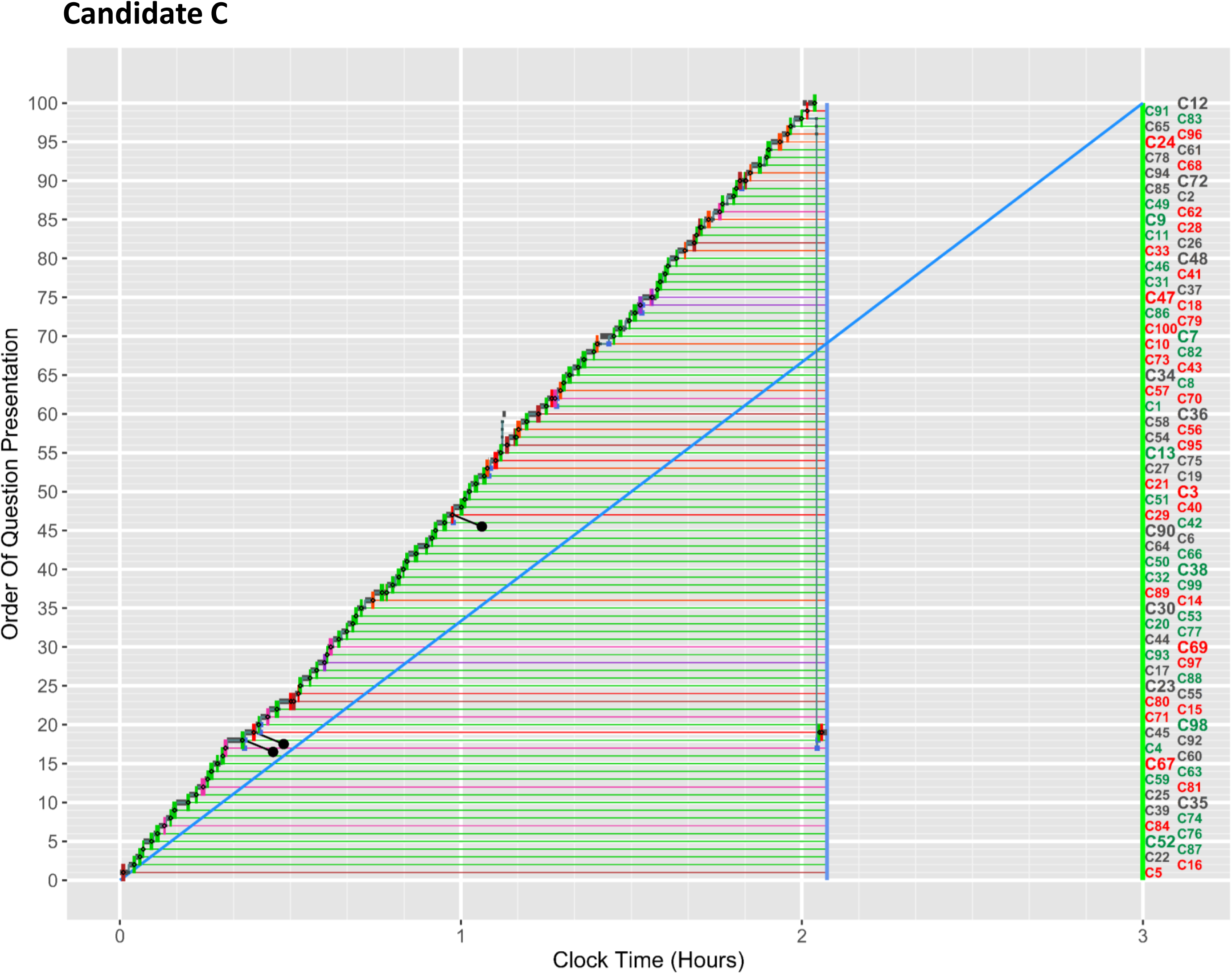
*Full ClickMap* for candidate C. See text for further details.

### Pinning or flagging questions

Not all questions necessarily need to be returned to, and the exam software allowed candidates to ‘flag’ or ‘pin’ items that they wanted to return to. Candidate D, in figure 6, is a high performing candidate, indicated by the large number of green answers, with the candidate’s performance above the 95^th^ percentile. On the main sequence, completed in about two hours ten minutes, twelve of the questions were flagged (indicated by the black pin pointing down to the right). The candidate then goes through all of the items once more in the second sequence, and in particular they carefully ‘unpin’ most of the questions pinned earlier, with unpinning shown by grey pins pointing down to the left.

**Figure 6:**
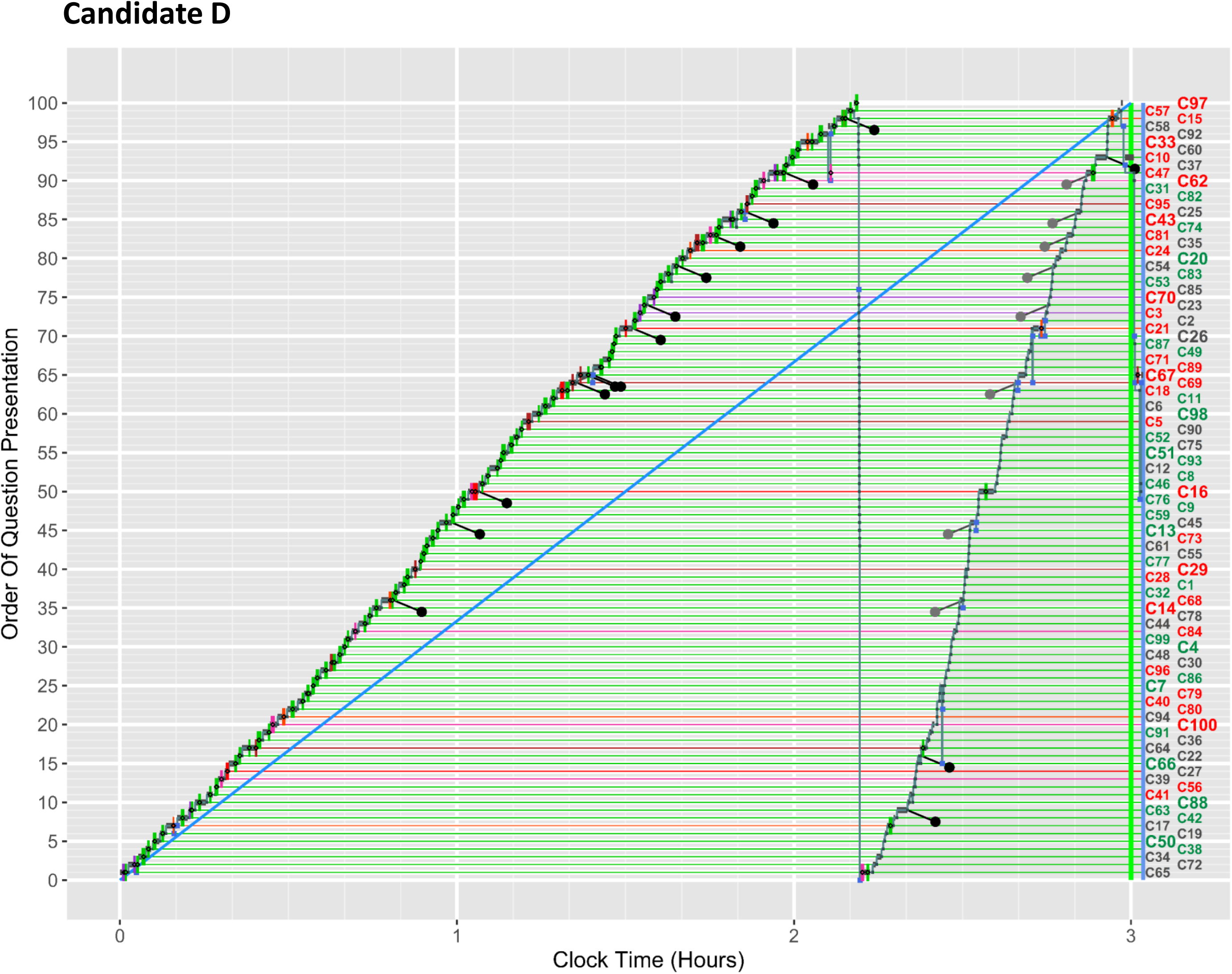
*Full ClickMap* for candidate D. See text for further details.

It is only of benefit to pin questions if sufficient time is left to return to them. Candidate E (figure 7) pinned eighteen items for review, but as can be seen by the deep ‘scalloping’ of the main sequence that early performance was slow, leaving insufficient time to return to the pinned items, and the last thirty items needing to be answered very quickly in the final half hour. Nevertheless, the candidate performed at about the 80^th^ percentile.

**Figure 7:**
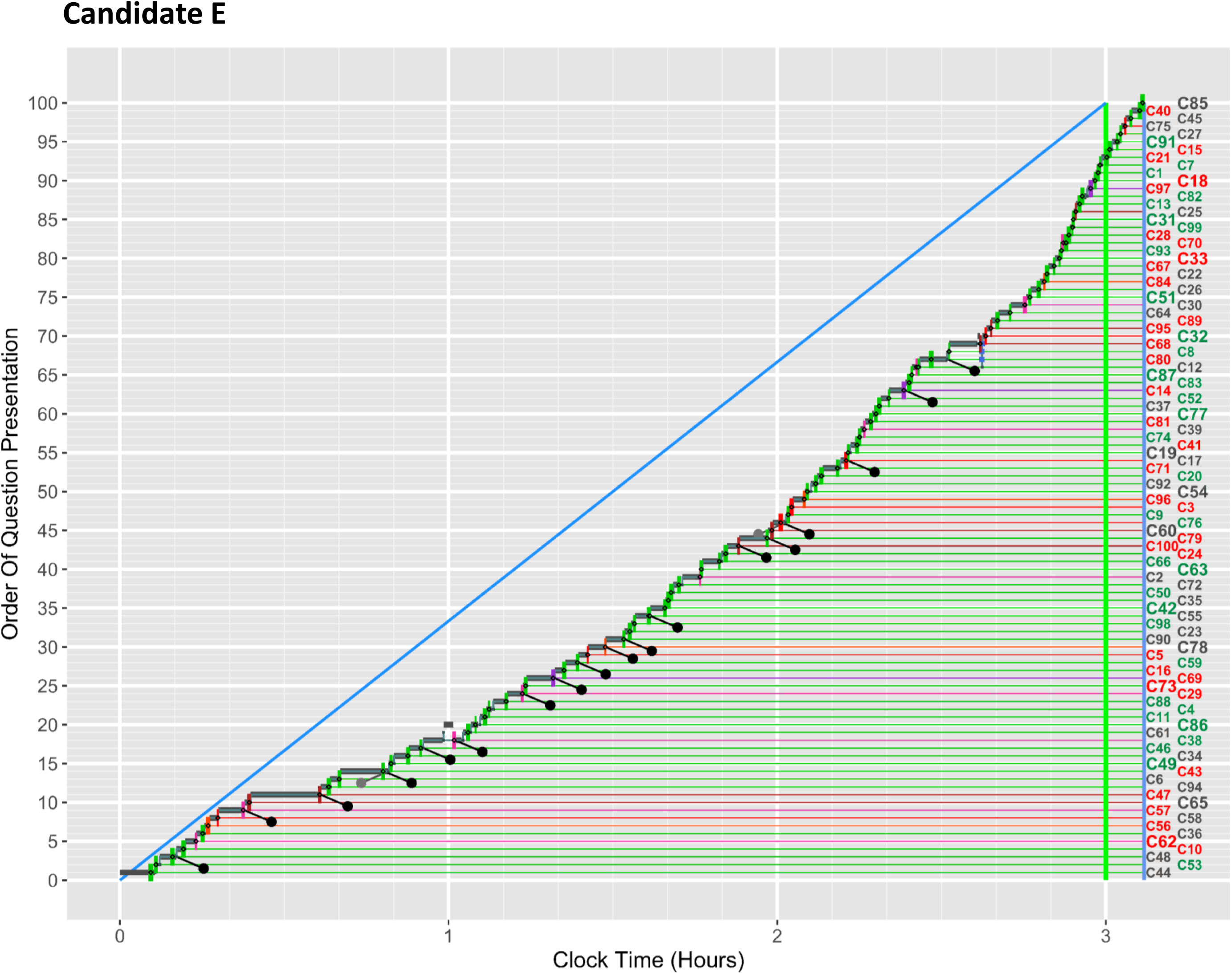
*Full ClickMap* for candidate E. See text for further details.

Some candidates review the entire paper on several occasions. Candidate F (figure 8) completed the main sequence in under one and a half hours, and in the process pinned over half of the items (52%) and then spent the next hour and a quarter on a second sequence, changing some answers, unpinning many of the pinned items, and adding a few new pins. Finally, in the last twenty minutes in a third sequence, many items were unpinned and a few answers changed. Candidate F performed at about the 85^th^ percentile.

**Figure 8:**
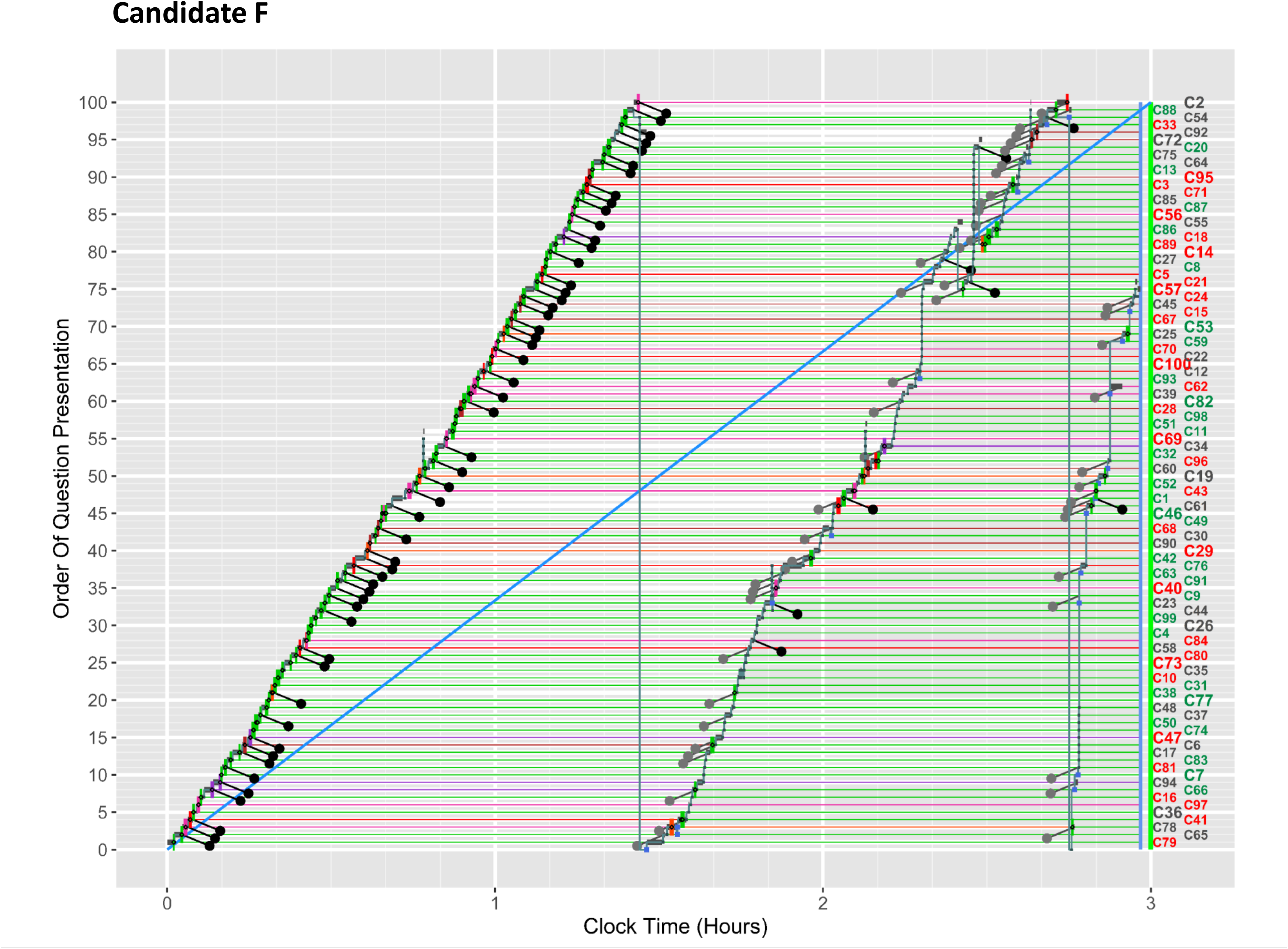
*Full ClickMap* for candidate F. See text for further details.

A very different strategy is shown by Candidate G (figure 9). The main sequence lasts only about 35 minutes, with only 22 questions answered, of which 21 (95%) were correct. The remaining 78 items are indicated in blue squares with white lines to indicate no answer was made at the time, the candidate moving instead to another question. The second sequence lasts about 55 minutes, with 33 of the 40 newly answered items being answered correctly (83%). A third sequence and then a brief fourth sequence follow, lasting 35 and 10 minutes, with only 9 of the 38 items answered correctly (24%). With answers now given to all items, a fifth sequence lasts 40 minutes, with three items having answers changed. The brief, sixth and final sequence lasts 10 minutes, with 87 items being rushed through at about six or seven seconds per item, as if something is being searched for, but the end result is that no changes made. Pins are used only twice, late in the exam, and the candidate mostly uses the system’s ability for noting items not answered to find items to be returned to; that usage is different to pinning which can flag already answered items. 95% of questions answered in the main sequence are answered correctly, with 83% in the second sequence, and 24% in the third and fourth sequences, showing that the candidate begins by picking off easy questions, leaving harder questions, and then the hardest questions, to return to, when they will have been viewed once or twice before. This is a sophisticated strategy overall, and its success is shown by the candidate being at about the 85^th^ percentile. There is a clear indication that the candidate is **well calibrated**, early answers mostly being correct, leaving items for which the candidate is less confident to later in the exam, and perhaps thereby giving more thinking time for the items. That thinking time may not only consist of actually looking directly at the question, but also having items ‘thought about in the back of the mind’, as it were.

**Figure 9:**
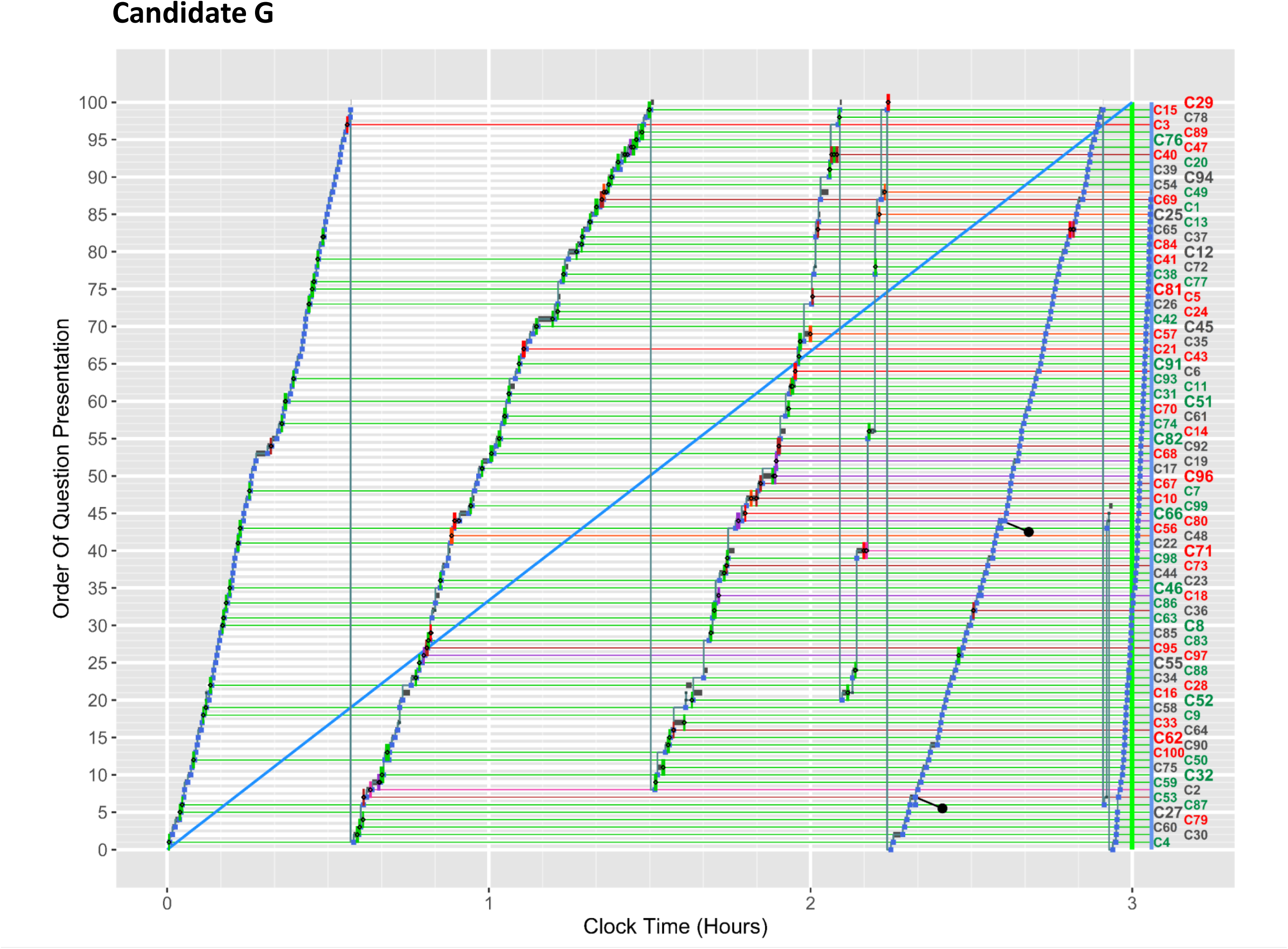
*Full ClickMap* for candidate G. See text for further details.

### Unusual ClickMaps

Candidate H in figure 10 is striking for the absence of any obvious main sequence. Items 1 to 9 are looked at, there is then a jump to item 18 and the candidate works backwards to item 10. By an analogy with genetic sequences, this can probably be called an **inversion**. A further jump goes forward to item 19 and the candidate works forward to item 25. A long jump then goes to the final item, 100, followed by working backwards to item 55, a jump further back to 38 and then a working forward to item 56. Again, thinking of genetics, this can be regarded as a **transposition**. By this point all 100 questions have not been covered, and it is possible that that fact is not very clear in the item display provided on the candidate’s screen. The candidate though returns to item 1, and works steadily forward to item 47, and only by about one and half hours into the exam have all of the questions been seen at least once. The main sequence has been chopped up into pieces and rearranged in forward and backward sections of inversions and transpositions. The remainder of the exam then shows various sections of ‘toing and froing’. Although the pattern looks somewhat chaotic, without talking to the candidate it is difficult to know the rationale for their behaviour, and the candidate did perform well at above the 80^th^ percentile.

**Figure 10:**
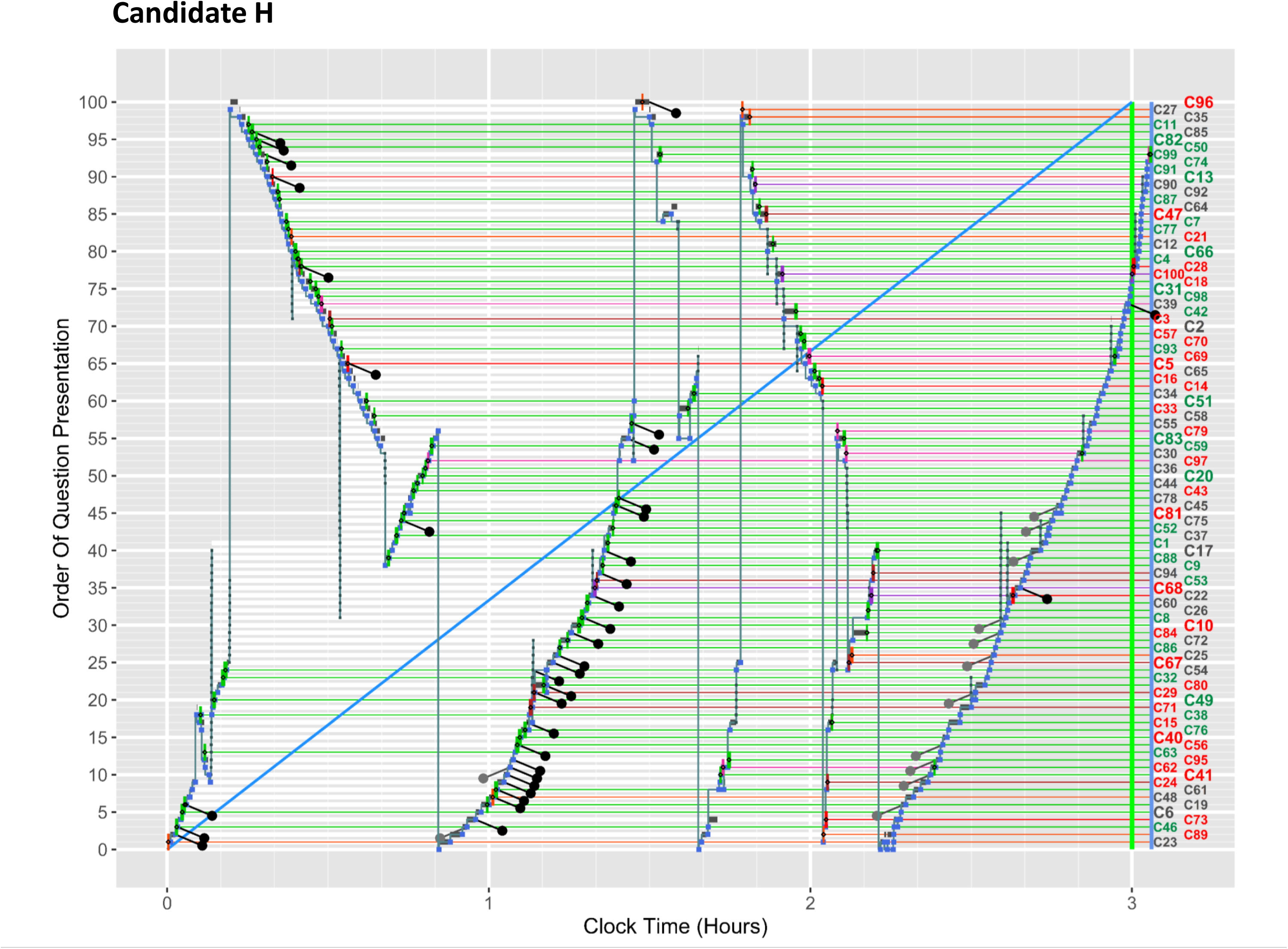
*Full ClickMap* for candidate H. See text for further details.

Candidate I in figure 11 shows a different type of jumping around. The main sequence is clear except for a brief inversion for the last eleven items. The curious feature is the seven vertical spikes, of varying heights, which are very brief with no item answered. These are probably produced by clicking quickly on the ‘up arrow’ button, followed by a jump back. The purpose of these is unclear, but they may be ‘horizon scanning’, so the candidate gets an idea of the questions ahead. The candidate performs above average at about the 75^th^ percentile.

**Figure 11:**
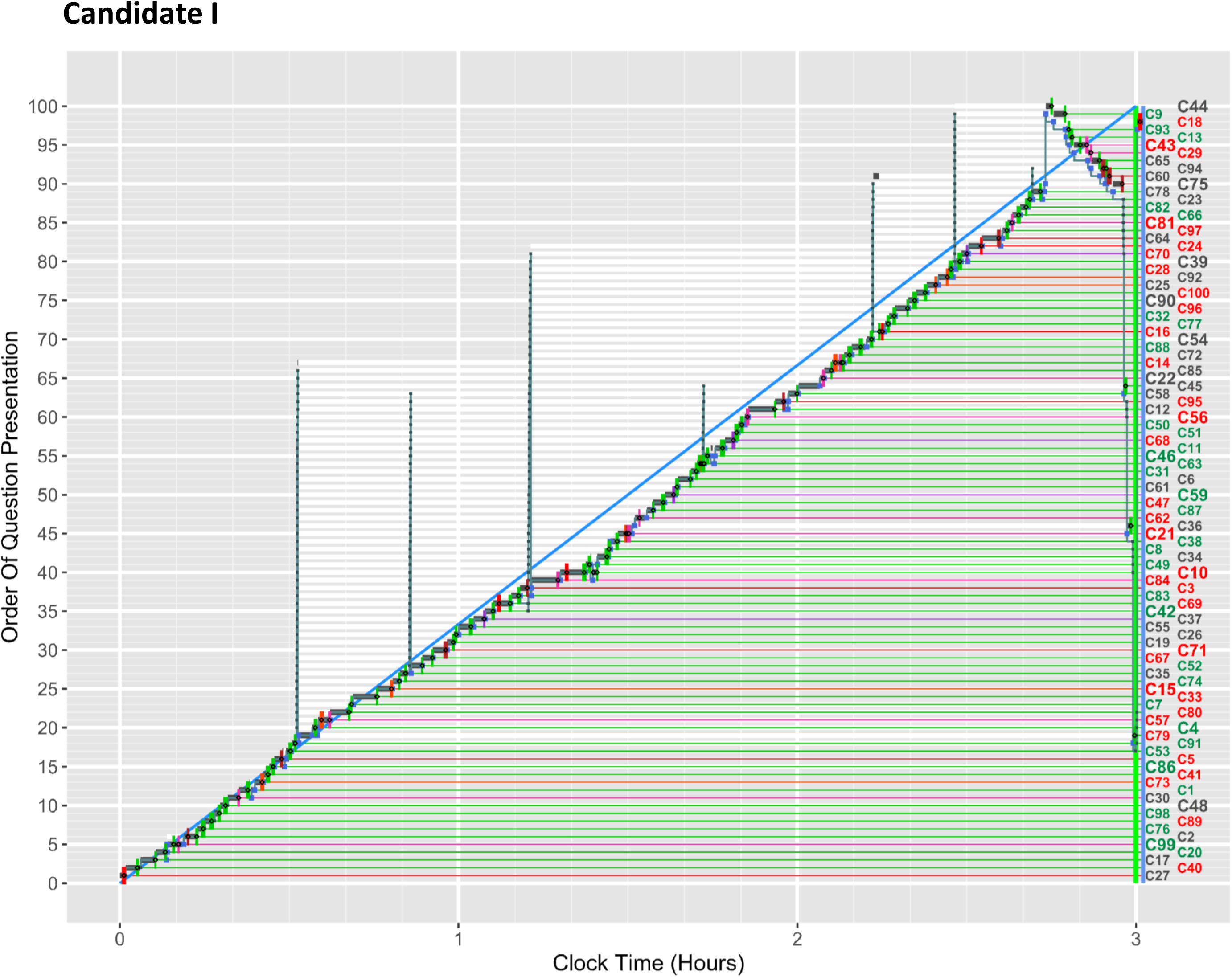
*Full ClickMap* for candidate I. See text for further details.

### Weak candidates

An obvious interest is in the performance of weak candidates with the possibility that inappropriate strategies may in part result in underperformance.

Candidate J in figure 12 was one of the dozen weakest candidates in the exam, and their poor performance can be seen by the excess of red over green lines, only about 45% of items being answered correctly. The *ClickMap* in figure 12 has features of figures 1 and 4. The main sequence finishes with a little time to spare, with answering being rather slow in the middle of the paper, but the timing then caught up in the last hour, leaving about ten minutes for revision. However, question 19, which had been looked at for a minute and a half on the first pass was still unanswered and had been left pinned. The candidate returns to that item for nearly ten minutes at the end of the exam; after several minutes they put in a wrong answer, followed by a different wrong answer, before finally entering the correct answer just before the exam finished. The candidate’s overall knowledge base presumably is weak, but their strategy seems systematic and careful.

**Figure 12:**
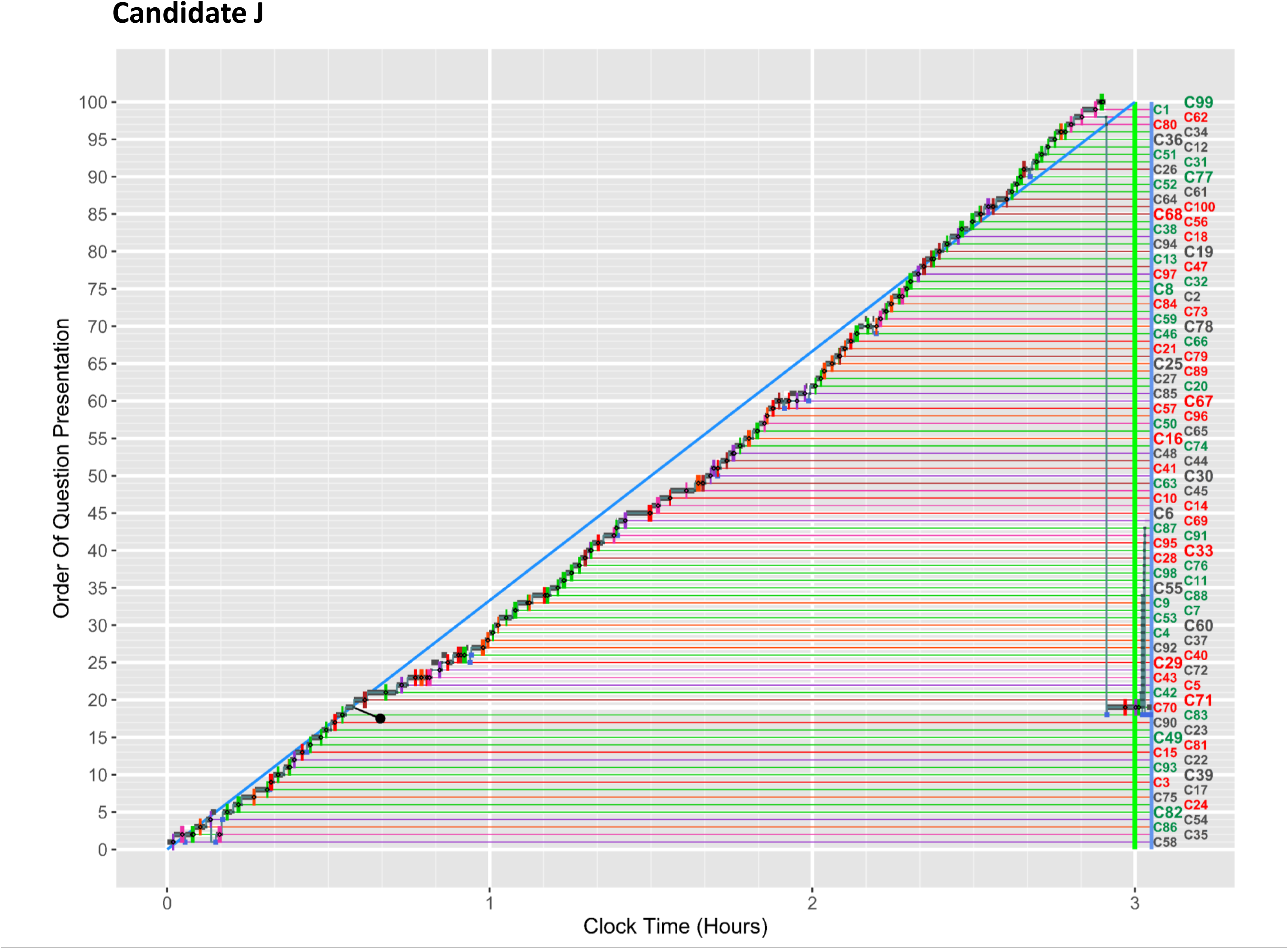
*Full ClickMap* for candidate J. See text for further details.

### Early and late changes

Candidate K in figure 13 was also one of the weakest candidates in the exam, with fewer than 50% correct answers, shown by the large number of red bars in figure 13. The main sequence finishes in about 2h:40m, leaving twenty minutes or so for returning to questions. The candidate had carefully pinned 19 of the questions, as can be seen in the *Full ClickMap* of figure 13 and the *Early ClickMap* of figure 14, with 13 pins later unpinned. During the revisiting of the pinned questions, there are six **late changes**, defined as changes occurring later than six minutes after the question has first been seen, and these are usually seen as part of a second or later sequences. Not so visible in figure 13, but clearer in figure 14, is the large number of **early** changes, as in Q4 where a wrong answer is chosen, a different wrong answer is then selected, and finally the original wrong answer is reselected. Overall 26 questions showed early changes in their answering. For comparison, candidate A made only 5 early changes, and no late changes. A host of interesting questions begins to emerge, as to the extent to which these differences between, say, candidates K and A partly explains the differences in their overall performance, with early changes perhaps indicating a lack of confidence or poor knowledge. Candidate K in figure 14 also has a different pattern of response, showing **non-answering periods**. Q35 is looked at for about 15 seconds, and then without answering, the candidate moves to Q36 for about 95 seconds, also without answering, and then returns to Q35 when after about 25 seconds the correct answer is given. The candidate then returns to Q36, and a gap can be seen in their responding in figure 14, and after nearly two minutes, a wrong answer is given. Such behaviour raises many questions.

**Figure 13:**
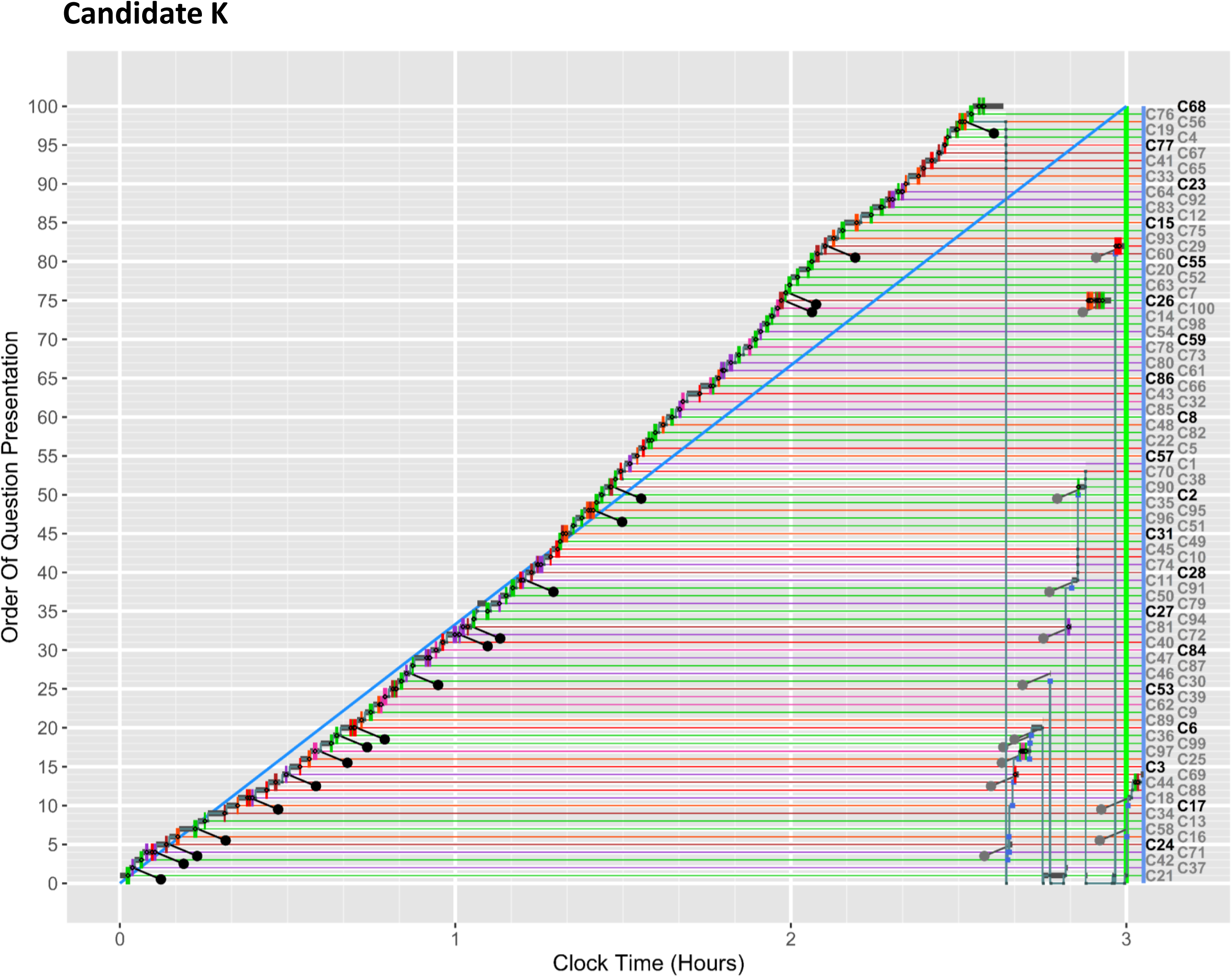
*Full ClickMap* for candidate K. See text for further details.

**Figure 14:**
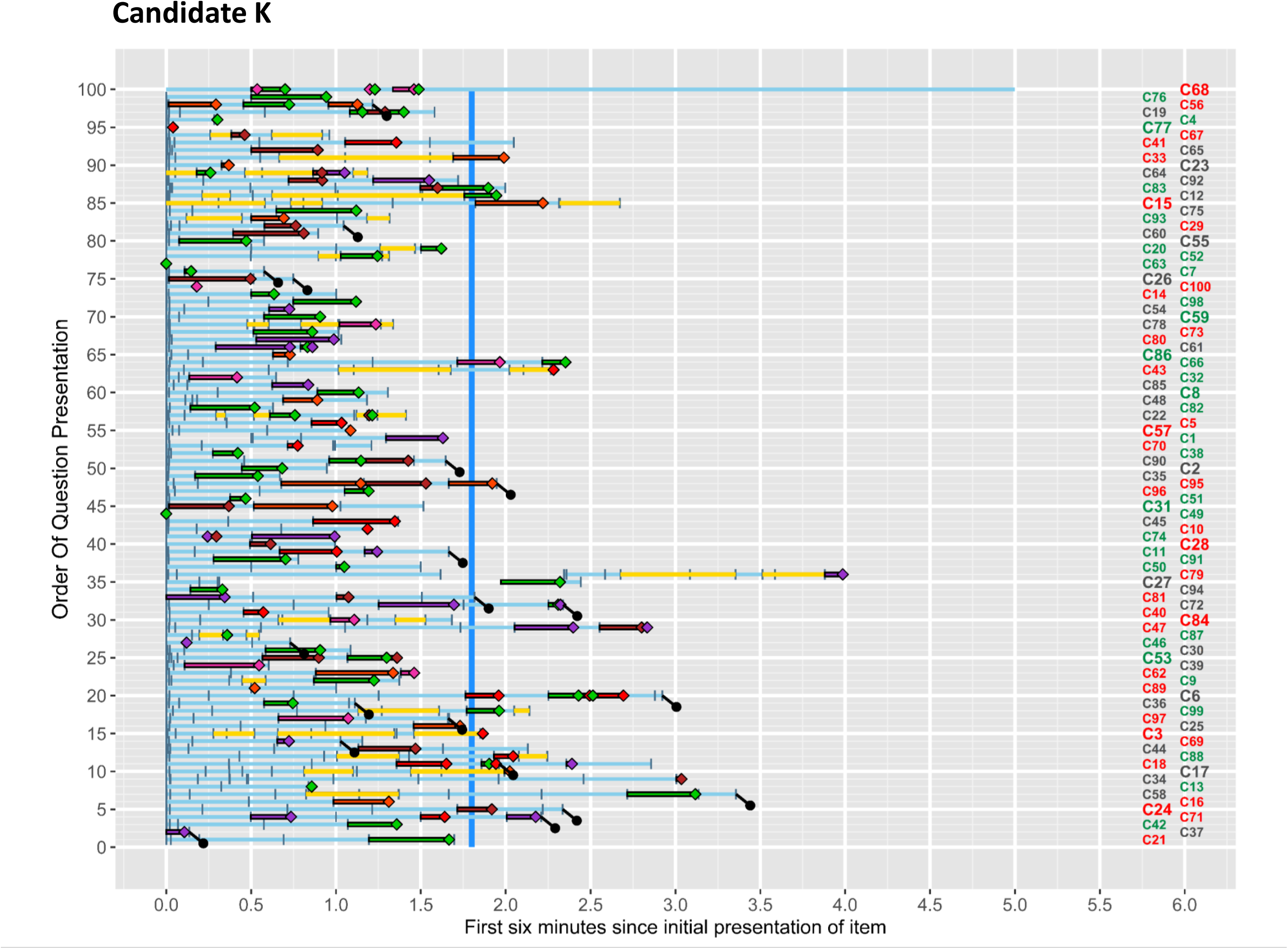
*Early ClickMap* for candidate K. See text for further details.

## Discussion

*ClickMaps*, the detailed plotting of what candidates are doing during every second of a three-hour exam, are characterised here in detail, for what we think is the first time. The *ClickMaps* shine a light on the complexities and subtleties of behaviour in examinations, and provide great potential for using this novel methodology to study how students interact with an examination which is delivered via computer. In particular, it is clear that candidates differ in their behaviour, which raises many questions on why candidates differ, and whether those differences matter in terms of examination performance, and possible feedback and counselling to candidates, particularly those who do not perform well.

The COVID-19 pandemic resulted in many undergraduate and postgraduate examinations moving, at speed, from traditional pencil-and-paper formats to online CBT formats, which provided a mass of information – the *ClickStream* -- about the moment-to-moment actions of the candidates as they navigated through the paper, chose answers and changed answers, returned to questions, looked at clinical images, and so on. No such information was available when the same exams had been carried out using printed question booklets, with answers indicated by making pencil marks in the pre-printed lozenges of optically scanned sheets. As mentioned earlier, ‘click’ refers generically to both mouse clicks, keyboard responses and navigation actions using fingers.

A *ClickStream* in its raw form is an undigested mass of numbers, typically provided in a CSV or similar format, comprising the many candidates and their multiple actions all in temporal order, with actions from different candidates interleaved. Few examiners are likely to be able to make sense of that torrent of numbers, with the result that the usual digested, summary output distributed to exam boards remains little different from that for pencil-and-paper exams, being a simple list of which items were answered correctly or incorrectly by each candidate. For an exam with 100 BoF questions, a pencil-and-paper exam for one candidate would provide just the 100 scores indicating the answers that had been given, whereas a *ClickStream* would contain perhaps 500 to 3000 or events per candidate, all time-stamped.

To our knowledge there is neither open-source nor proprietary software available for generating *ClickMaps*, but it is possible to use programs such as *R* [14], as we have done here, to create *ClickMaps*, although the programming effort is non-trivial. We hope in the future to create a proper *R* package for analysing *ClickStreams* and generating *ClickMaps*, but that is not straightforward because different online exam providers use different data formats and different codes for events, not all providers providing all types of information, such as when a candidate merely looks at a question without answering it, or is viewing an associated image.

### Reading ClickMaps

*ClickMaps*, like many of the images used in science, not only summarise large amounts of data, but also present those data in ways which are readily comprehensible to the human mind, brains being poor at processing numbers *en masse* but being very good at understanding complex images. Data visualisation is an art as much as a science, but modern computer graphics software allows the creation of high quality images which when well devised can exploit principles of good graphical design [17-19] and neural, perceptual and cognitive processing [20]. Well-constructed graphs, which often present data using Gestalt principles, with colours, shapes, and other visual features for grouping and separating different image components, are often understood very rapidly, as geographical maps in atlases have learned how to do over last half of a millennium. Medical images of various sorts can also be equally effective. Our experience of *ClickMaps* is that clinicians rapidly treat them like X-rays or MRI scans, understanding them very quickly, and learning to compare and contrast *ClickMaps*, and make inferences about the underlying candidate behaviour.

### How long does it take to answer a question?

Although not the main purpose of the present report, an interesting question is why some questions take longer to answer than others, and the rich data in a *ClickStream* provides much information, as was shown in 2001 using a USMLE assessment [9]. At least three factors are probably involved; some questions are intrinsically easier or harder than others; some candidates know more or less than others; and some questions happen to be placed later in the exam where there is less time remaining to answer them, and there is also less time for revisiting questions. Plotting such factors together is not easy, because the graph would be multidimensional.

### Developing and extending *ClickMap displays*

As with any mapping process, maps can be developed and extended to include further information. The *ClickMaps* in figures 1 to 14 are functional and easy to understand, but developments are still being considered. At present questions are presented in presentation order, but that makes it hard to compare items on, say, their overall difficulty, since the hardest item will be at any of 100 different positions across the different candidates. It is also possible to plot *Early ClickMaps* sorted not by presentational order but by order of average candidate difficulty, perhaps with the distribution of response times of candidates overall also indicated, so that candidates can see whether they are particularly having problems with easier or harder items. No doubt there are many other aspects of candidate behaviour that can also be plotted, and these need exploring. The present analysis has only scratched the surface of the richness of the questions that can be asked.

### The vicissitudes of random question ordering

Randomness of presentation order means that all candidates have an equal chance of seeing each question at a particular position in the exam. That though does not entirely guarantee that all things are equal for all candidates. Candidate A for instance, in figure 1, started with two difficult questions (C81 and C96), and their first relatively easy question was only reached when they encountered C50 in the seventh position. In contrast, candidate D in figure 6 had five easier items and five moderate items in the first ten, only encountering a difficult item at the 11^th^ question. It is possible that starting the exam with a string of easy or a string of difficult questions may lead candidates to believe that the exam is particularly easy or particularly hard and alter subsequent behaviour, with a ‘set’ towards choosing easier or harder responses. Considering the other end of an exam, candidate A had two of the easier questions presented as the 99^th^ and 100^th^ questions, when time was running out for them, and they needed to answer quickly. Overall all such things will balance out across candidates, but individual candidates may not perhaps see it in that way. Candidates who left more time for returning to questions would have fewer problems with difficult questions suddenly appearing at the end of the exam.

### Classifying *ClickMaps*

*ClickMaps* clearly differ visually even when looked at on a computer screen as ‘thumbnails’, suggesting large-scale, macro-differences in approach. However, zooming in on the *ClickMaps* also shows micro-differences, as in the comparison of the *Early ClickMaps* of candidates A and K in figures 3 and 14. A challenge is to produce a classification of *ClickMaps*, which might consider such features as the extent of the main sequence and how soon it is finished, the presence of secondary and other sequences, the use of flagging or pinning of items to be returned to, and so on. Although that could be done on an *ad hoc* basis, it might also be that machine learning approaches to visual image classification could be successful. Given that large postgraduate examinations may have several thousand candidates, a machine-based approach might be helpful either for research purposes or for directed counselling. A formal nomenclature would be useful, and we cannot help noticing that many of the events shown in the *Full ClickMaps* are similar to those described in genetic data, including duplications, inversions, deletions and transpositions. There are also similarities to the behaviour of organisms in Skinner boxes on fixed-interval schedules which show ‘scalloping’ [21] and other patterns. Whether the similarity is formal or functional is unclear, but scalloping can occur in cumulative response records of human behaviour where there are strong time constraints [22-24].

### Individual differences in exam strategies

The *ClickMaps* we have shown for candidates A to K make clear that candidates can have quite different strategies and approaches. That raises a host of educational and cognitive questions which can only briefly be mentioned here. We assume that strategic differences are relatively stable, and we hope to confirm that in the future. Motivations and approaches for studying and practising medicine are varied. Undergraduates often have *surface*, *deep* or *strategic* (*achieving*) motivations for studying medicine, and as a result adopt different study habits and processes, which as postgraduates can extend into different approaches to work (*surface disorganised*, *surface rational*, and *deep*) [25]. It seems likely that such differences may well affect how candidates prepare for and also how they take examinations. We also note that, in medical students, previous exam failure can result in a shift to maladaptive surface-learning approaches [26], with similar processes potentially occurring after repeated failure in postgraduate examinations.

Other influences on the origins of strategic differences are less obvious; are they rationally decided upon, or are they perhaps idiosyncratic responses to previous successes and failures in exams (and medical students and doctors can be seen as professional exam takers)? Do strategies which work at one academic level, such as for medical students in their early years, where rote learning can sometimes be effective, work as well in later years or for postgraduate assessments, where responses at a deeper cognitive level are required? How does aptitude, and also personality, affect the approach to exams? We cannot answer those questions here, but hope to look at them in the future.

A student on a graduate-entry undergraduate course told one of us of, “that her approach to answering SBAs is to do the first 10-25, then go to the last 10-25 and do those. She repeats this pattern and meets in the middle. Her thinking is that she doesn’t like having the weight of 200+ questions in front of her and finds it more psychologically pleasing to chip away at both ends”. However, “She knows it’s all randomised and so there is no external advantage to it”. How common such perception and reasons are requires further study.

### Candidates with reasonable adjustments

Although not described here, a proportion of candidates have reasonable adjustments and, as a result, are allowed extra time for taking exams. How such time is used is very unclear, as is the extent to which these candidates behave differently when answering questions. The issue is particularly pertinent in candidates with dyslexia, where 25% extra time is often routinely allowed, but it is unclear whether or when a benefit actually occurs (and certainly not all candidates allowed extra time seem always to use it).

### Changing Answers

Many, but not all, candidates, change answers, with some vacillation back and forth between chosen alternatives as seen for example in candidates A and K. Such behaviour may be even more extensive than that explicitly recorded in *ClickMaps*, since back in 1928, not long after MCQs were developed, it was pointed out, “it is … probable students change their minds one or more times *prior to making any mark at all*” [27][our emphasis], and of course such changes are invisible to researchers. Changed answers in *ClickStreams* must always therefore be an under-estimate of true uncertainty. In the late 1920s it was also noted that first impressions are more likely to be wrong than are changed answers [28]. Nevertheless many students and teachers in the 1920s suggested that *first* answers are more likely to be correct than changed answers. Nearly 50 years later, in 1977 Stoffer et al could still refer to the “stable effect” whereby changed answers are more likely to be correct, despite a “stable misconception” that it is thought better *not* to change answers, a conclusion also reached elsewhere [29]. A modern version of the erroneous belief in first answers may derive from claims for the efficacy of thin-sliced, fast thinking rather than thick-sliced, slow thinking [30, 31]. A 1994 meta-analysis confirmed that changed answers in MCQs are more likely to be correct [32], an effect seen in a more recent study [33]. We also have *ClickStream* data from a recent undergraduate and a recent postgraduate exam showing that changed answers are more likely to be correct. Of greater interest and novelty for present purposes is that it is yet unclear whether benefits arise both from early changes and late changes to answers, or whether these differ in their benefit, and that will be analysed in a future paper. *ClickMaps* and *ClickStreams* provide large datasets which should allow greater investigation of the details of when there is a benefit from changing answers. Overall there is a sense in which not changing answers, or in not leaving time to review and to make such changes, may be seen as a maladaptive strategy.

### Candidate insight, confidence and metacognitions

Cognitive tasks of any type can be carried out correctly or incorrectly, and MCQ examinations are no exceptions, answers being right or wrong. Candidates often have intuitions or insights about the correctness or otherwise of an answer varying from a feeling of complete certainty, through to seeing it as “a total guess”. Such *metacognitions* themselves vary in accuracy, resulting sometimes in dangerous situations of high confidence in inaccurate information, or accurate knowledge which is ignored due to low confidence. Once a question is answered then further metacognitions appear, “allowing us”, as Stephen Fleming says, “to reflect on whether we might be wrong and perhaps to change our minds or alter our response” [34]. Experimental studies can measure confidence and hence metacognitions directly [35], and can also assess their neural underpinnings [36]. Examinations typically cannot record confidence ratings, although higher confidence can often be inferred indirectly for a question answered quickly, with a single answer, and no return to the question for re-inspection. Accurate metacognitions in exams may also have a more general importance to clinical practice, since metacognitive accuracy can generalise across tasks [37].

### Maladaptive strategies

Whether some *ClickMaps* indicate maladaptive strategies for taking CBT exams is unclear, but it would certainly seem that running out of time and then having to answer many questions in the final few minutes, or even not answering all of the questions, are not good ways to take an exam. Whether other strategies are also maladaptive is less clear, but vaguely remembering an earlier question on, say, hyperparathyroidism, and then at the end of the exam blindly searching through the hundred or so question texts for that condition may not be a good use of time left at the end of a test (and we note that software of exam providers often have no text search function). It is probably for that reason that a majority of candidates seem to take surprisingly detailed notes on the blank paper with which they are provided (and those notes will be part of a study to be reported later). If some strategies are less optimal than others, particularly if candidates do not realise that to be the case, then there may be a justification for intervention and remediation. Given that almost all studies show that candidates benefit from changes made to answers, then the large proportion of candidates who show the patterns typified by candidates A or C, who either leave no time to revisit questions, or who choose not to do so despite having time available, may suggest there might be benefit from feedback and greater insight.

### Visual questions

The *Early ClickMaps* in figures 3 and 14 show how candidates switch between windows when answering questions with visual material, having to alternate between reading question text and then looking in detail at visual images. It seems likely that candidates may well differ in their strategies when answering such questions, particularly when they return to them, putting a load on verbal and visual working memory, which probably differ in capacity between candidates [38]. Exam software may also hinder some candidates if there is a requirement, as here, to toggle back and forth between image and question text, or in some cases to scroll up and down, rather than allowing the two to be seen on screen simultaneously.

### Candidate responses to ClickMaps

Intuitively it seems possible that candidates themselves might wish to know more about their own *ClickMaps*; and in most assessments there is usually a demand by candidates for more feedback on their performance. Whether candidates would recognise their own maps in comparison with others, and how they would explain and make sense of those maps could form the basis of an interesting qualitative study.

### Cheating, collusion, and prior knowledge

An issue for all high-stakes exams is that some candidates may cheat [39], and while that is relatively easy to control in invigilated examination halls, the opportunities for cheating and collusion have probably increased with the advent of remotely proctored examinations often sat in insecure locations such as candidates’ homes [40]. Although we will not discuss the issue in detail here, the rich data from the *ClickStream* allows some unusual patterns of behaviour to be spotted more easily. In ***Bivariate ClickMaps***, which we have described elsewhere[41], the clock times for responding to questions are plotted on the horizontal (x) axis for one candidate and on the vertical (y) axis for a second candidate. Randomised question order means that generally two candidates should not be answering the same canonical question simultaneously. If however candidates navigate to the same canonical question at the same time then collusion is possible [41]. ‘Question hopping’ to result in concurrent viewing of questions is unusual and potentially suspicious behaviour [40]. Likewise very short response times for correctly answered items, especially but not always, during concurrent viewing may suggest suspicious anomalies in responding [42]. A separate type of cheating involves ‘candidate prior knowledge of examination content’ [43] or ‘item preknowledge’ [44], where exam content has leaked to groups of candidates, a signature for which is a group of candidates giving unusually rapid correct answers for difficult items, and which can be detected using data from the *ClickStream*, and displayed on variant *ClickMaps*.

### Modelling response processes

The present work was stimulated by various discussions not about whether candidates get answers right or wrong, but about the cognitive processes by which they arrive at right or wrong answers, and why some questions are harder than others. *ClickStreams* give detailed information on the timing of answers in computer-based tests, but further analysis probably requires a formal model of how decisions are made. Although most medical assessments are ‘best-of-five’, most cognitive modelling considers two-choice alternatives, and the timing of the decisions that are made [45, 46], although such models can be extended for more than two choices [47]. In psychology a typical two-choice-alternative study asks a participant to make a judgement between two texts or events, A and B, with prior probabilities usually set at 0.5. As the experiment proceeds a participant acquires further information, and at some point sufficient evidence is accumulated in favour of, say, A (or B), a response threshold is reached, and A (or B) is given as the answer. Such ‘diffusion models’ can also be applied in decision-making studies. Such experiments are typically not time-limited. Although the tasks are more complicated, MCQ questions behave in a formally similar way. However multiple-choice exams usually have time limits, so that a decision has to be made not only for which answer is more likely to be correct, but also whether there will be an opportunity cost for waiting longer to make a decision as time spent on one question reduces the time available for answering other questions. A candidate is therefore collecting more evidence as time passes but also racing against the clock [46]. A key finding from looking at the *ClickMaps* is the sometimes exquisite control of timing shown by the candidates, answering the 100 questions very precisely within the three hours, sometimes also leaving sufficient time for answers to be revisited. Managing time in a three-hour exam is itself a complex cognitive process and *ClickMaps* provide a rich context for studying and understanding time management. Formal modelling using the rich timing data available in the *ClickStream* is clearly required.

### Strategy differences or differences in knowledge base

Candidates might perform poorly either if they have a poor strategy, or they have a poor knowledge base for the material in an exam. Separating out those reasons for poor exam performance, as for instance with candidates J and K who are both weak candidates is important if counselling of weaker candidates is to take place successfully. More research is clearly needed on this important issue. A weak knowledge base may be indicated by alternating between answers, or answering rapidly but with a wrong answer (particularly with a rare wrong answer which most other candidates do not consider), or by pinning items suggesting a lack of confidence in answers provided. A weak knowledge base is probably also indicated by wrong and slower answers to items that most candidates answer correctly and quickly.

## Conclusions

*ClickMaps* represent a novel, intuitive, and visually powerful tool to interrogate how students interact with an online single-best-answer format examination. For the first time insight is gained into the detailed temporal dynamics of the cognitive processes going on in candidates’ minds when interacting with assessment material. The methodology opens the door to a gold mine of information that has potential to strengthen our understanding of student ability, develop evidence-based guidance on examination strategy, and radically evolve how we provide granular feedback on exam performance.

## List of abbreviations

BoF: Best of Five question
BMJ: [was] British Medical Journal; now is just the proper name of the journal.
CBT: Computer-based testing
ECG: Electrocardiogram
MCQ: Multiple Choice Question
MRCP(UK): Membership of the Royal Colleges of Physicians of the United Kingdom
PNG: Portable Network Graphic file
SBA: Single Best Answer question
USMLE: United States Medical Licensing Examination
VSA: Very short answer question

## Supporting information

Supplementary image files

## Data Availability

Data from MRCP(UK) examinations are confidential and not available.

## Declarations

### Ethics approval

This paper was discussed at the meeting of the MRCP(UK)’s Management and Policy Board meeting of 23^rd^ February 2023. The Board approved the plan to publish the paper and agreed that ethical approval was not required as it was based on an audit of data rather than research. The Board noted that the decision tool of the Health Research Authority (HRA) confirmed that the study was audit and not research, and therefore did not require ethical approval (https://www.hra-decisiontools.org.uk/research/question1.html).

### Consent for publication

Not applicable.

### Availability of data and materials

Data from MRCP(UK) examinations are confidential and not available.

## Funding

No external funding was provided to support this research.

### Authors’ contributions

All authors are involved in running, administering and researching into computer-based testing, either at undergraduate or postgraduate level, and all had separately explored the analysis of CBTs, and discussed together the issues involved. The software for the *ClickMaps* program shown here was written by ICM, who also wrote the first draft of the paper. All authors contributed to revisions of the paper, and all have approved the final draft.

## Acknowledgments

We are grateful to staff at MRCP(UK) Central Office who have assisted in importing the *ClickStream* data and interpreting the various results.

## Additional files

There are 22 high-resolution files for the *Full ClickMaps* and *Early ClickMaps* for candidates A to K, named as **Candidate_n_Full_ClickMap.png** and **Candidate_n_Early_ClickMap.png**, where **n** is the Candidate identification from A to K. All files have been anonymised by removing the top rows of the images, so that candidate code number etc have been removed. Images are .png files and are therefore compressed in a lossless way without loss of resolution. They can be viewed in any standard photo software, including Photos on PC, and also Mozilla Firefox. In general the default for viewing .png files should work. The high resolution should allow zooming into details as required.

Additional File 1: Candidate_A_Full_ClickMap.png

Additional File 2: Candidate_A_Early_ClickMap.png

Additional File 3: Candidate_B_Full_ClickMap.png

Additional File 4: Candidate_B_Early_ClickMap.png

Additional File 5: Candidate_C_Full_ClickMap.png

Additional File 6: Candidate_C_Early_ClickMap.png

Additional File 7: Candidate_D_Full_ClickMap.png

Additional File 8: Candidate_D_Early_ClickMap.png

Additional File 9: Candidate_E_Full_ClickMap.png

Additional File 10: Candidate_E_Early_ClickMap.png

Additional File 11: Candidate_F_Full_ClickMap.png

Additional File 12: Candidate_F_Early_ClickMap.png

Additional File 13: Candidate_G_Full_ClickMap.png

Additional File 14: Candidate_G_Early_ClickMap.png

Additional File 15: Candidate_H_Full_ClickMap.png

Additional File 16: Candidate_H_Early_ClickMap.png

Additional File 17: Candidate_I_Full_ClickMap.png

Additional File 18: Candidate_I_Early_ClickMap.png

Additional File 19: Candidate_J_Full_ClickMap.png

Additional File 20: Candidate_J_Early_ClickMap.png

Additional File 21: Candidate_K_Full_ClickMap.png

Additional File 22: Candidate_K_Early_ClickMap.png

