## Supplementary figures and images for "Visualising candidate behaviour in computer-based testing: Using *ClickMaps* for exploring *ClickStreams* in undergraduate and postgraduate medical examinations"

### Candidate_A_Early_ClickPlot.png

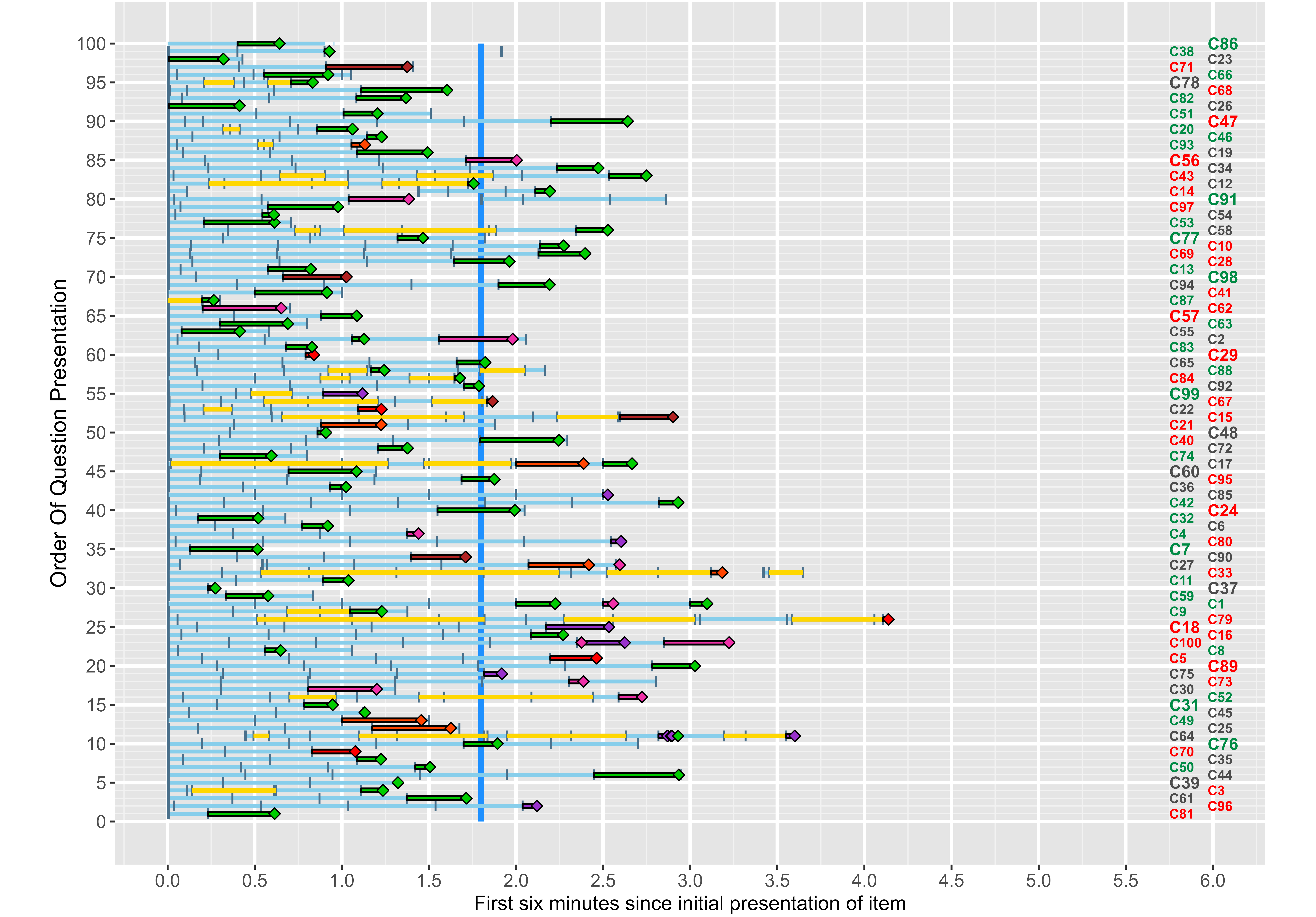

### Candidate_A_Full_ClickPlot.png

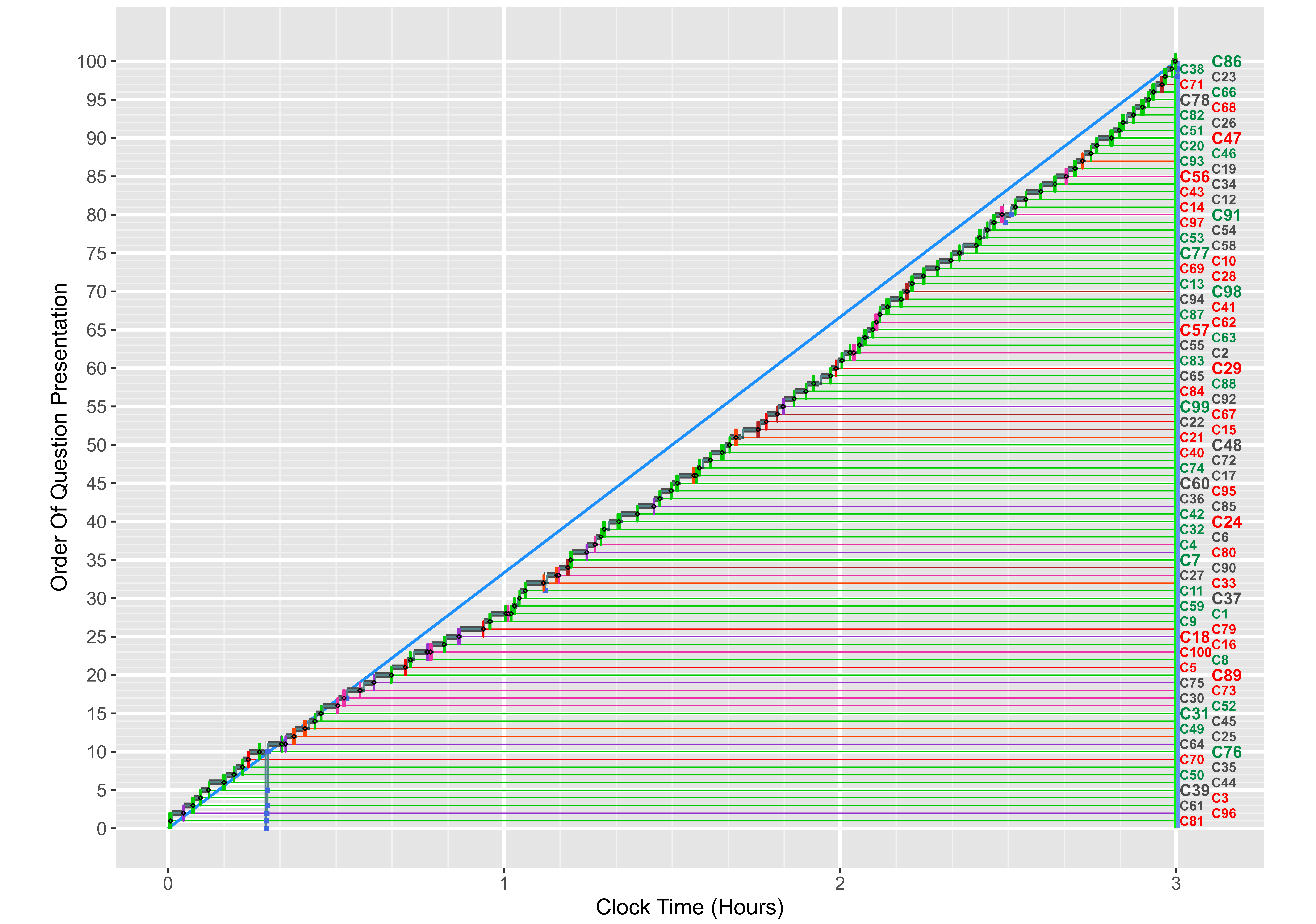

### Candidate_B_Early_ClickPlot.png

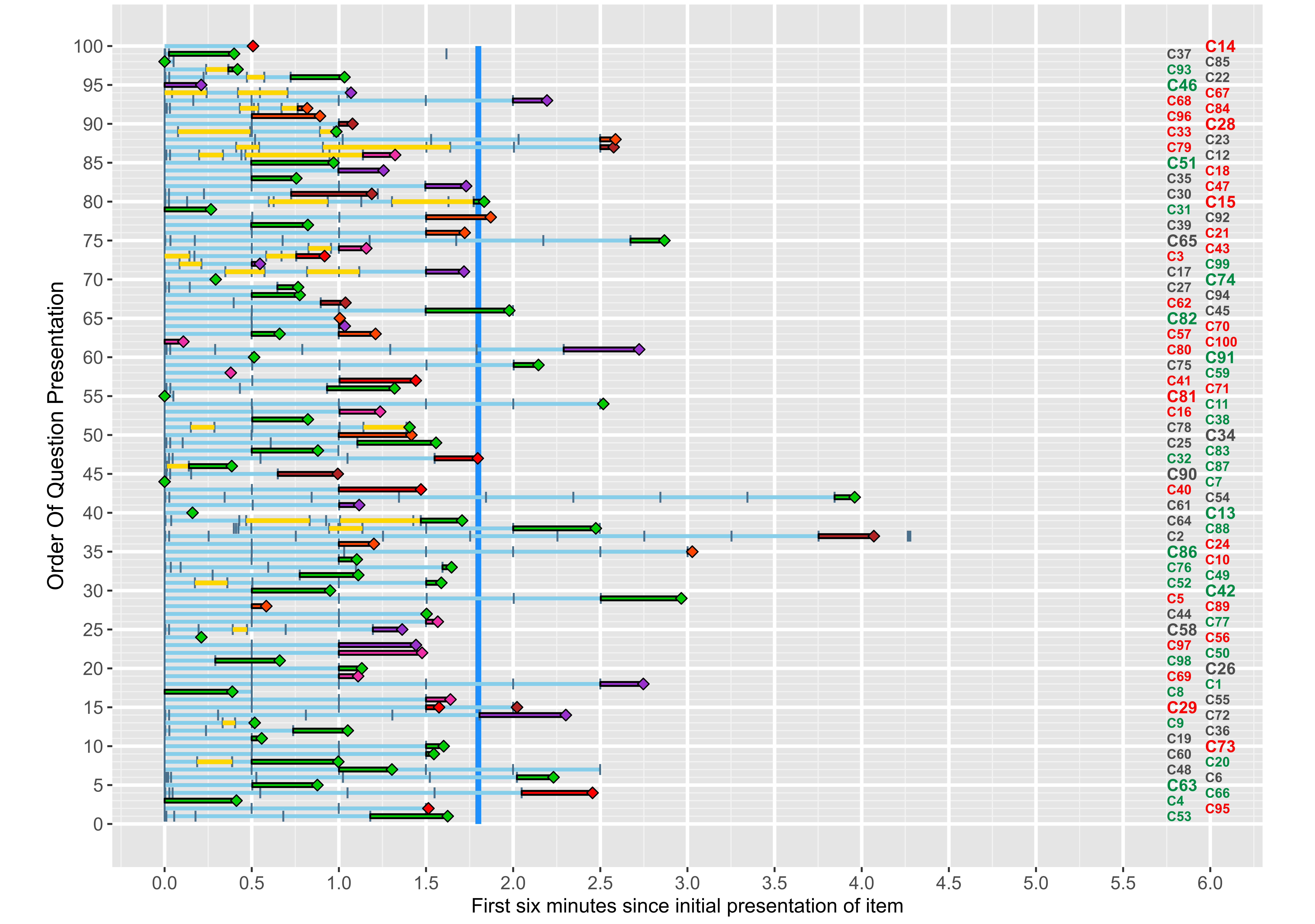

### Candidate_B_Full_ClickPlot.png

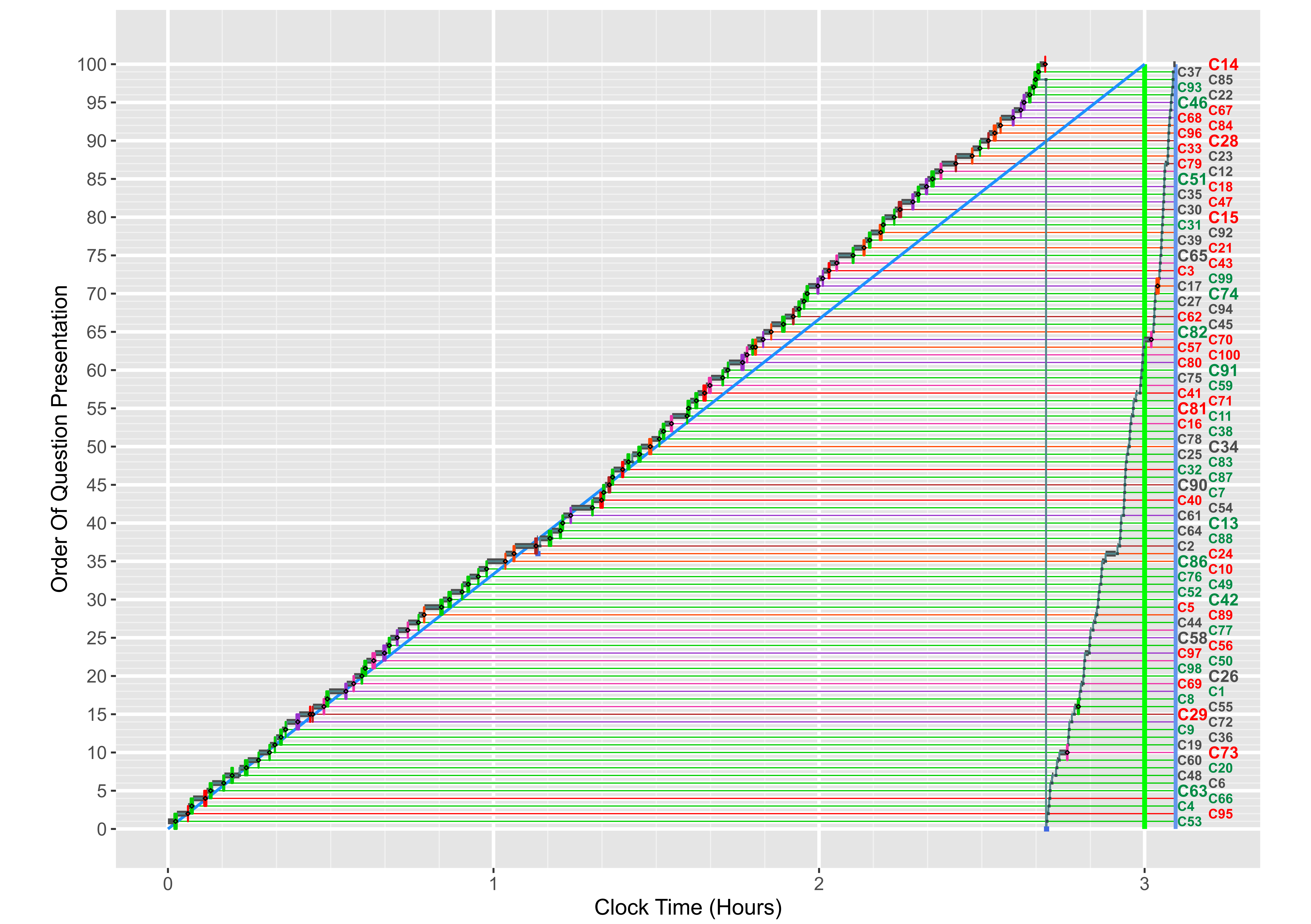

### Candidate_C_Early_ClickPlot.png

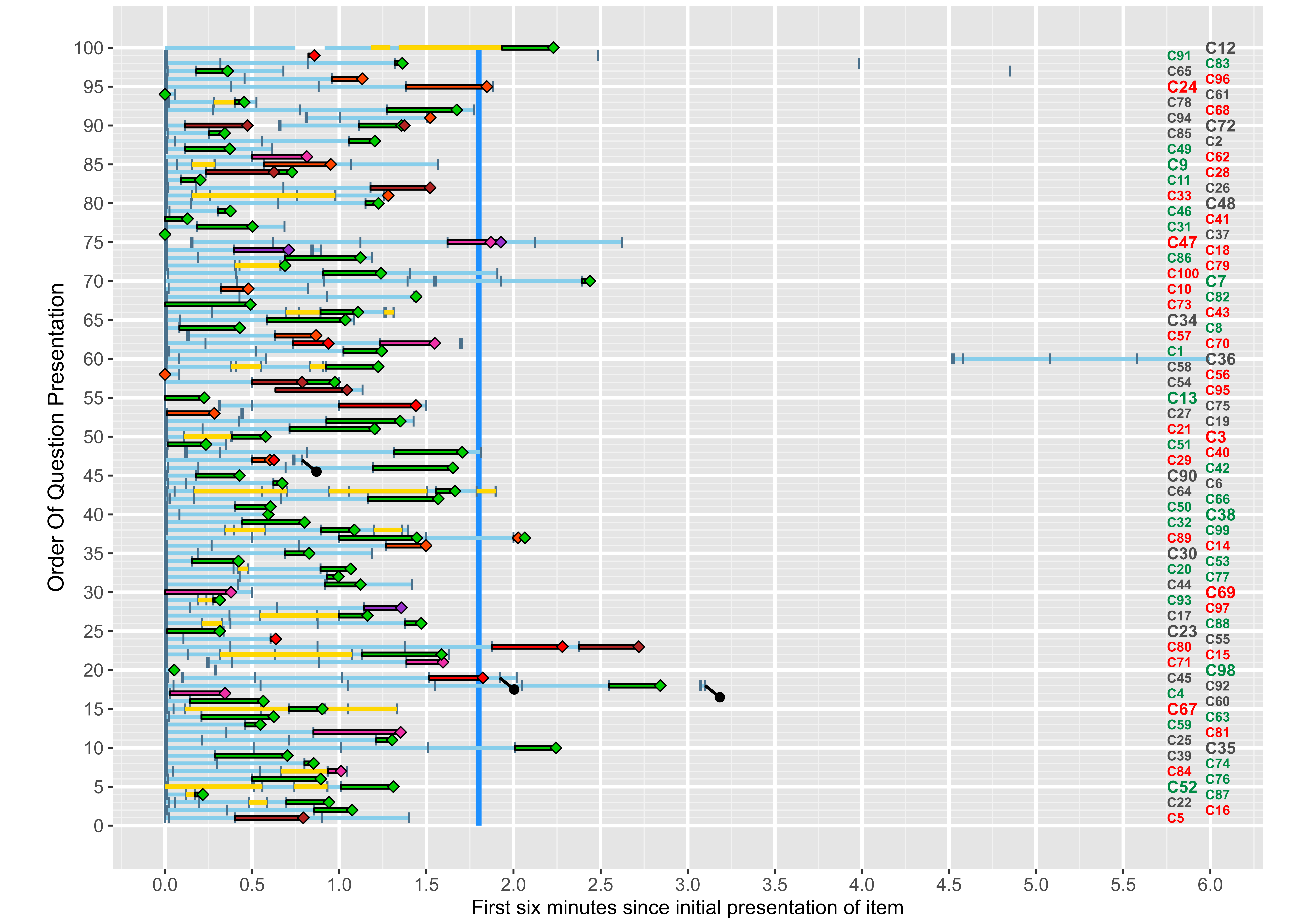

### Candidate_C_Full_ClickPlot.png

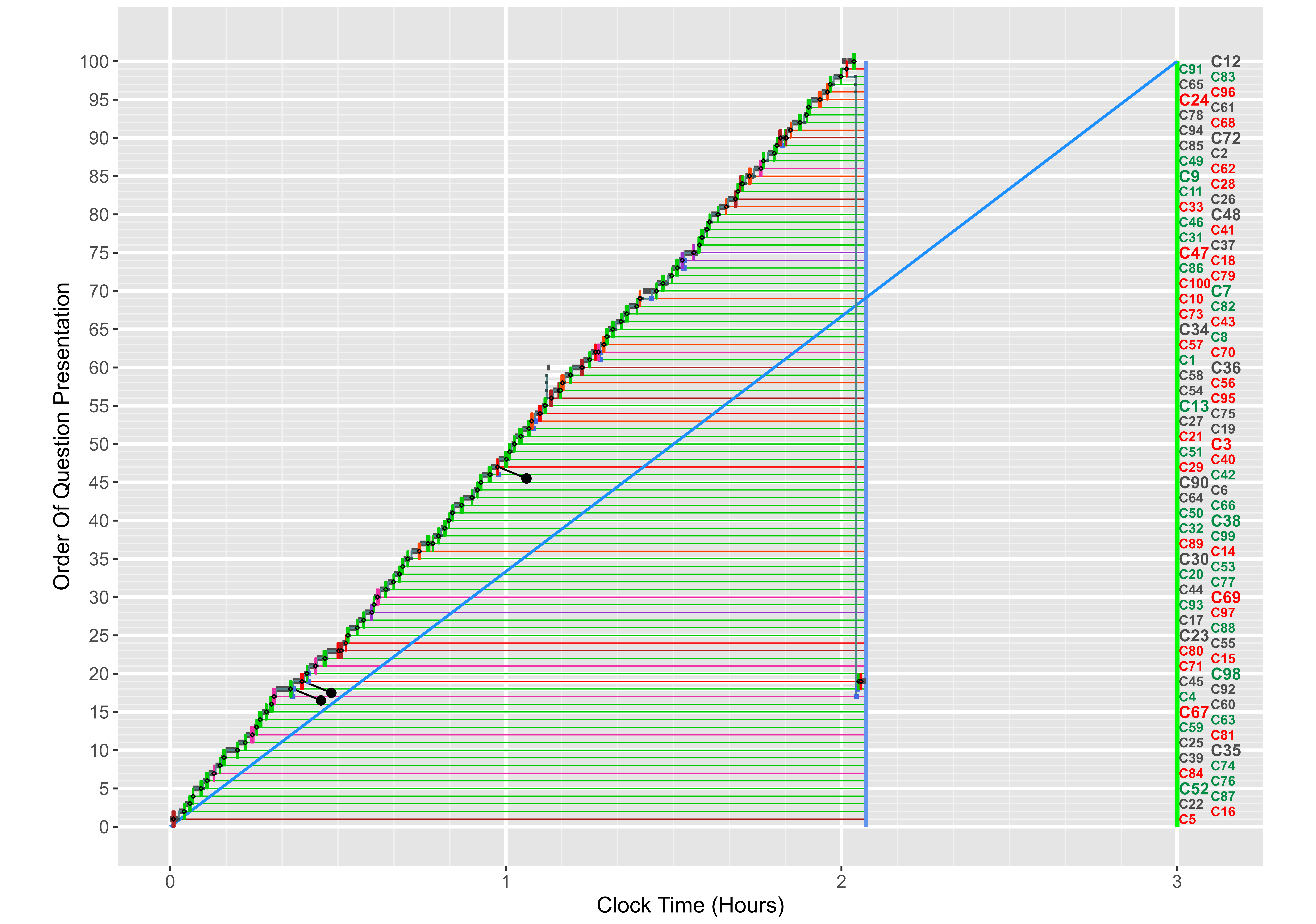

### Candidate_D_Early_ClickPlot.png

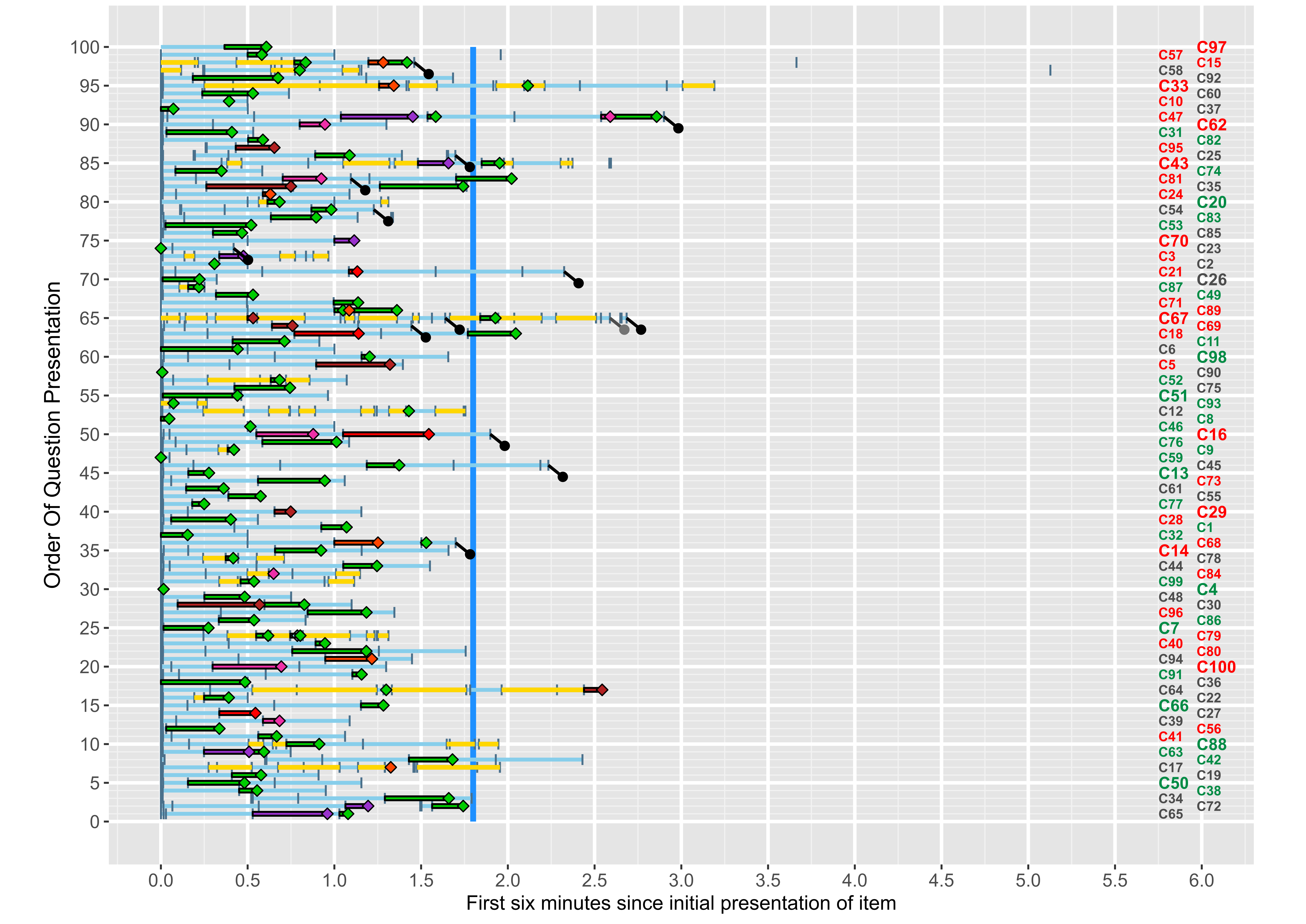

### Candidate_D_Full_ClickPlot.png

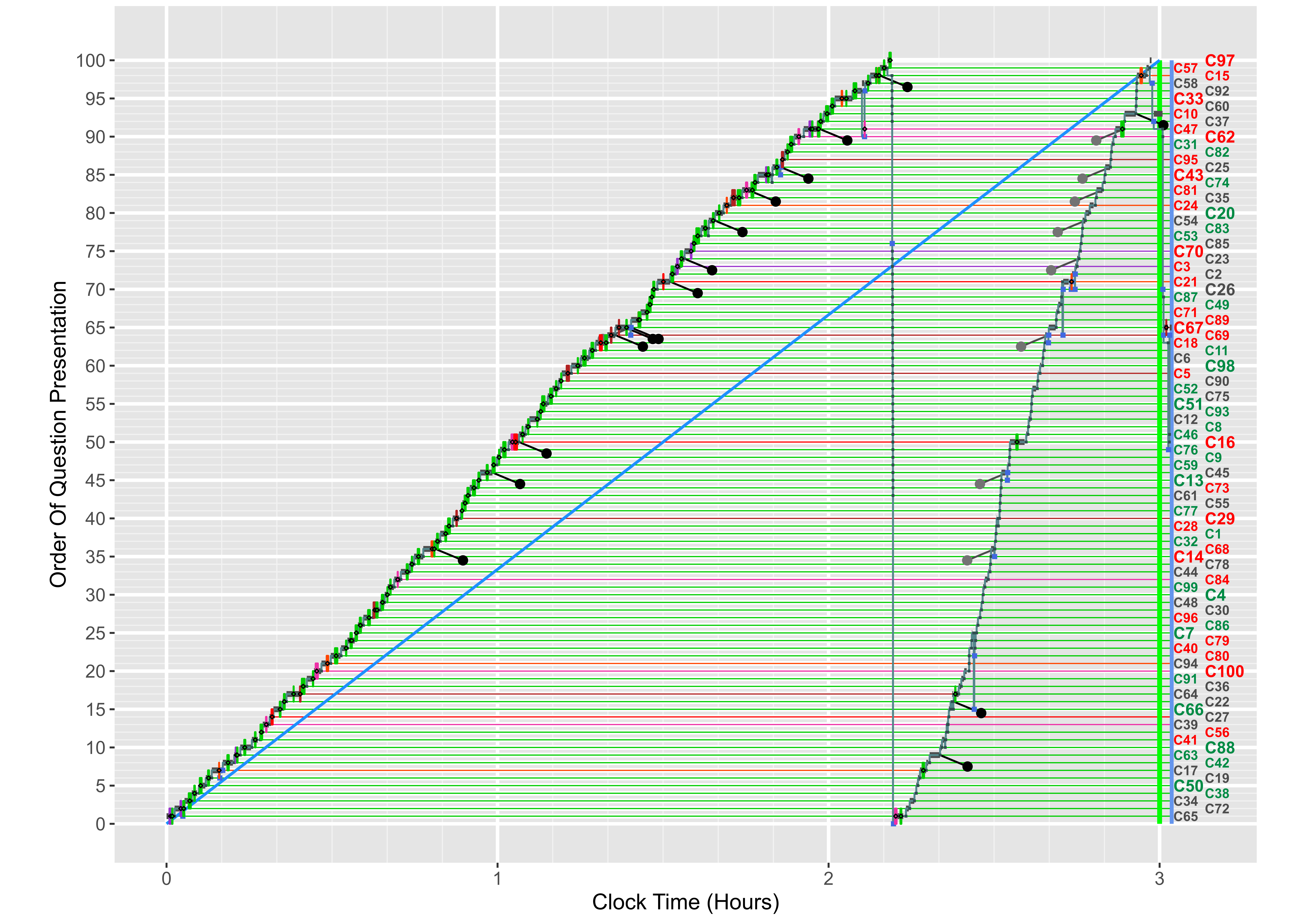

### Candidate_E_Early_ClickPlot.png

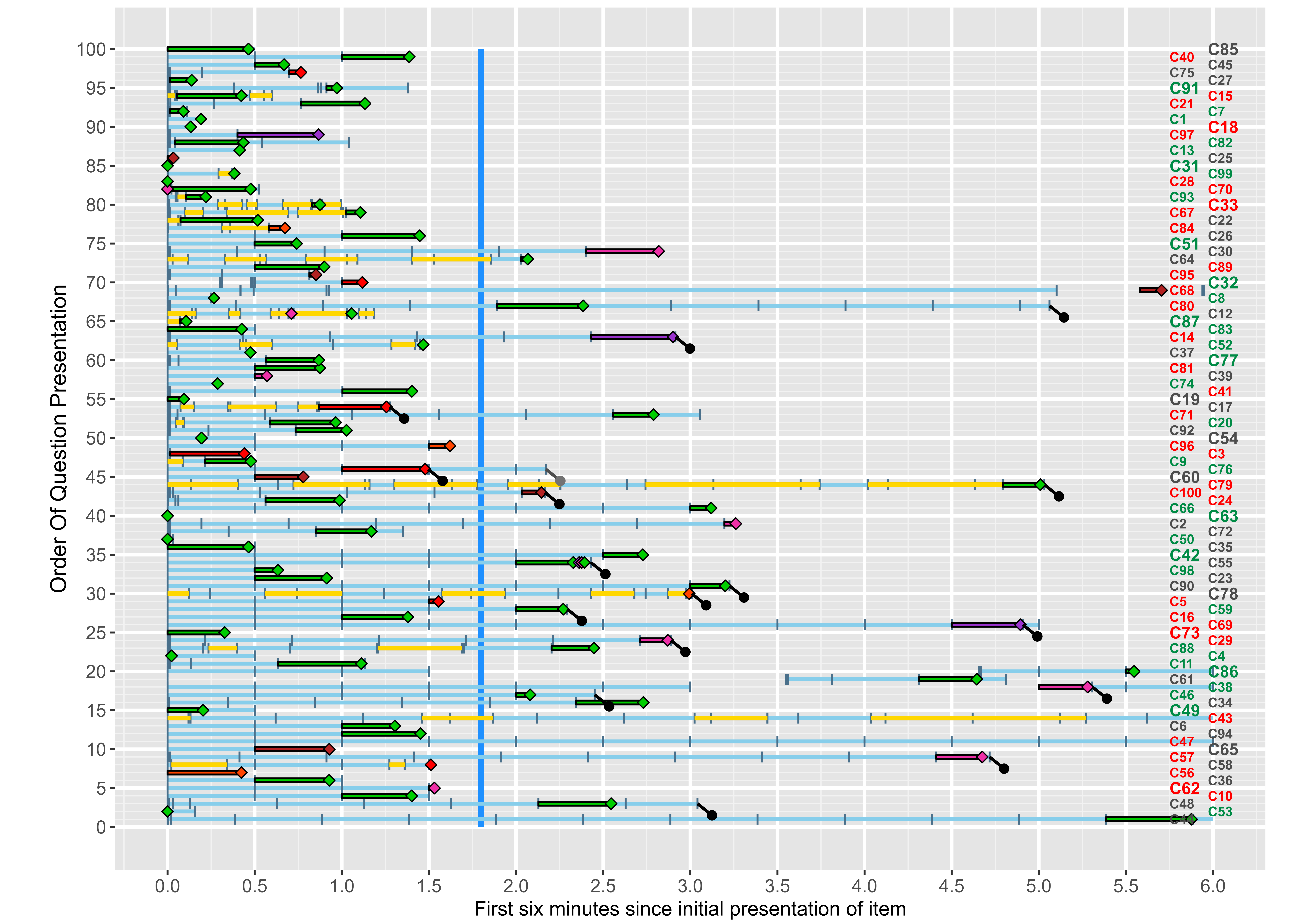

### Candidate_E_Full_ClickPlot.png

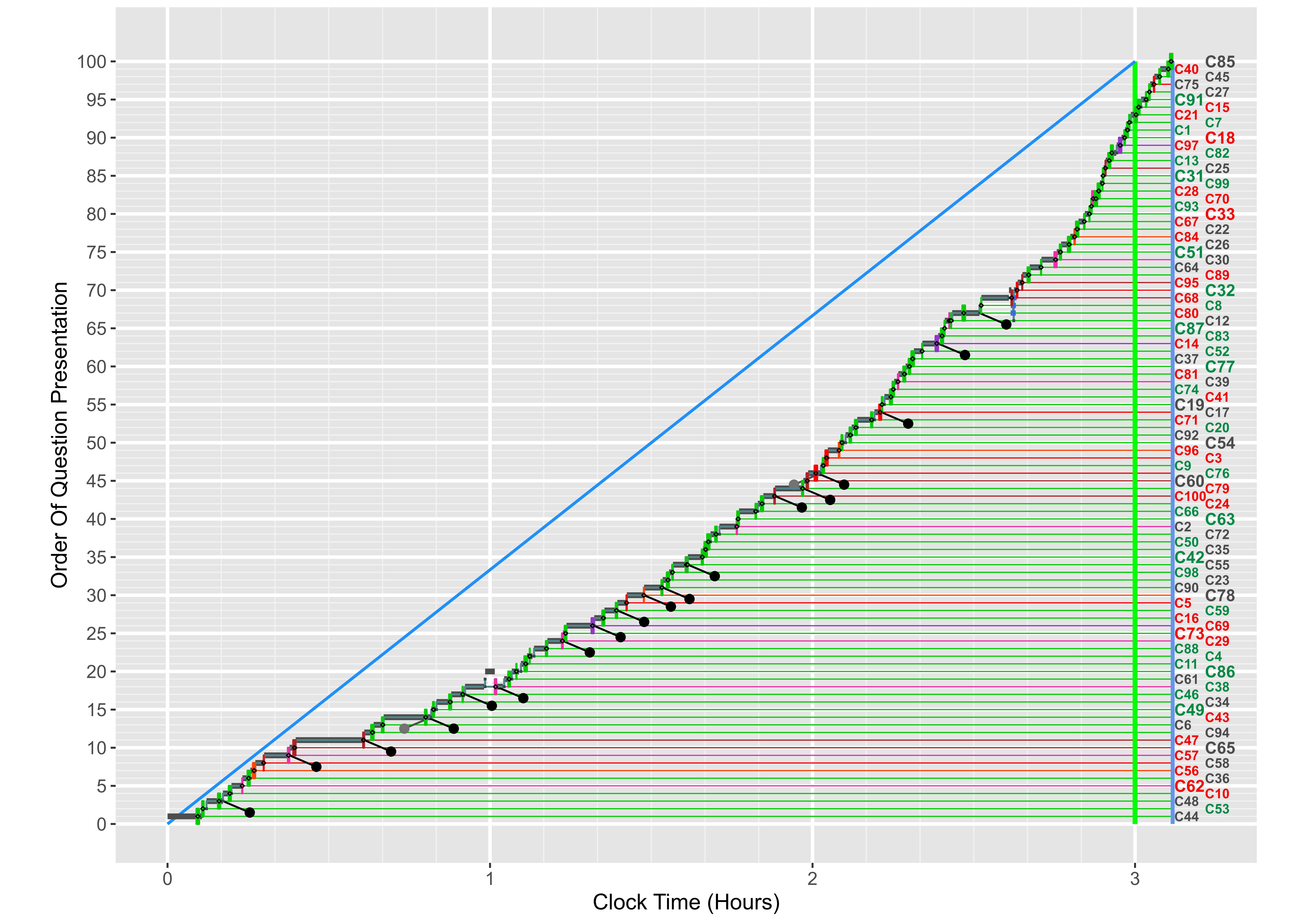

### Candidate_F_Early_ClickPlot.png

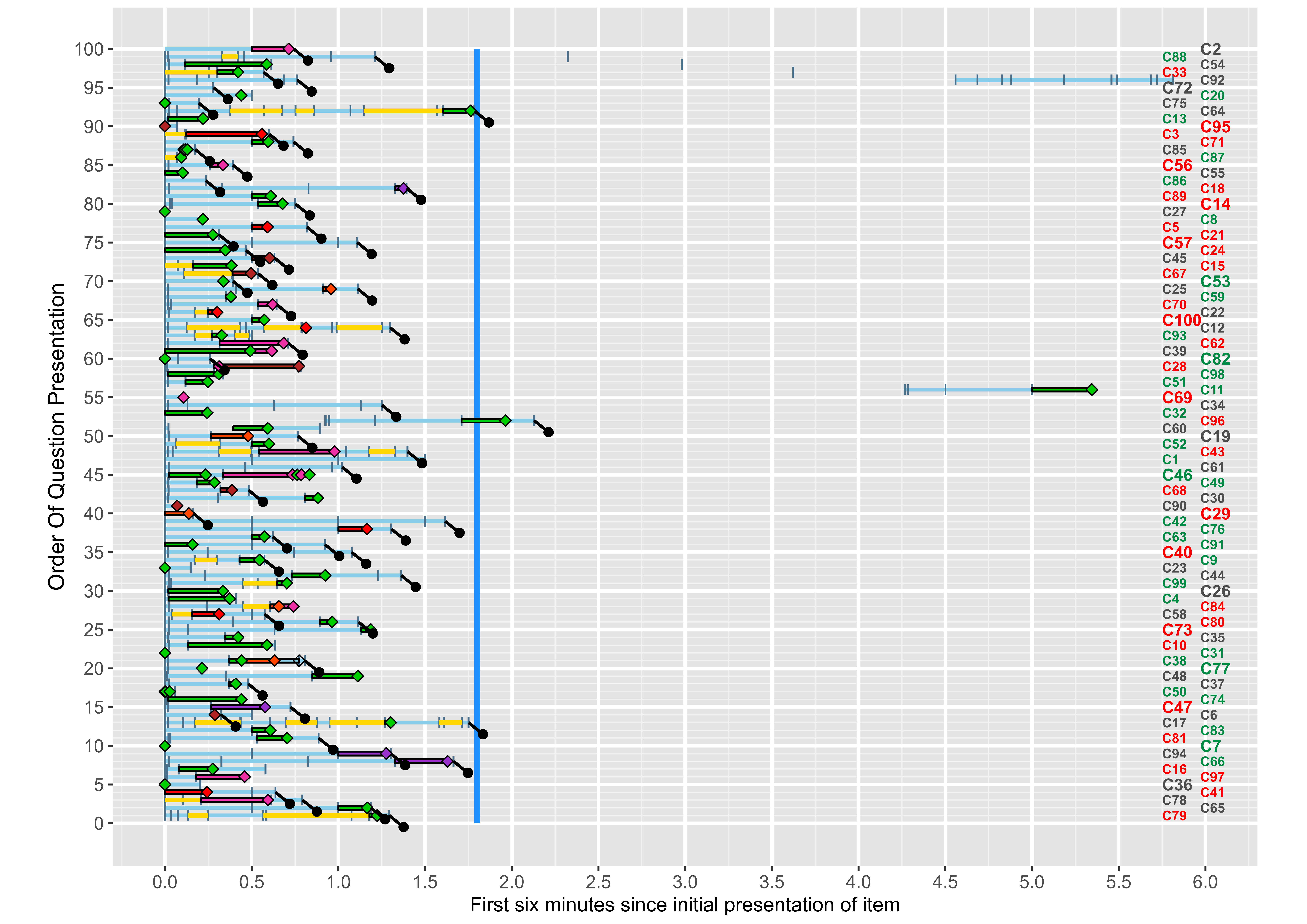

### Candidate_F_Full_ClickPlot.png

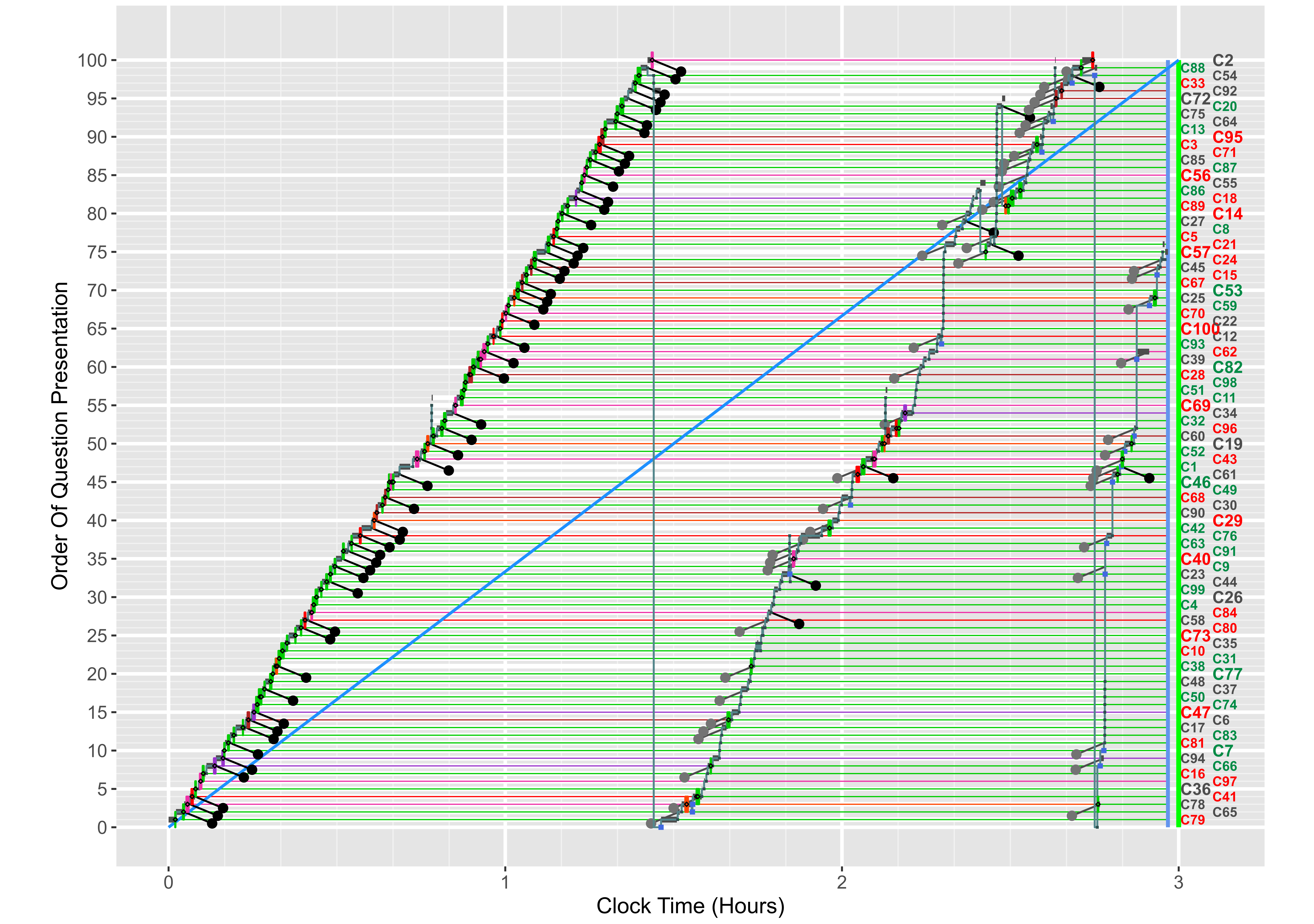

### Candidate_G_Early_ClickPlot.png

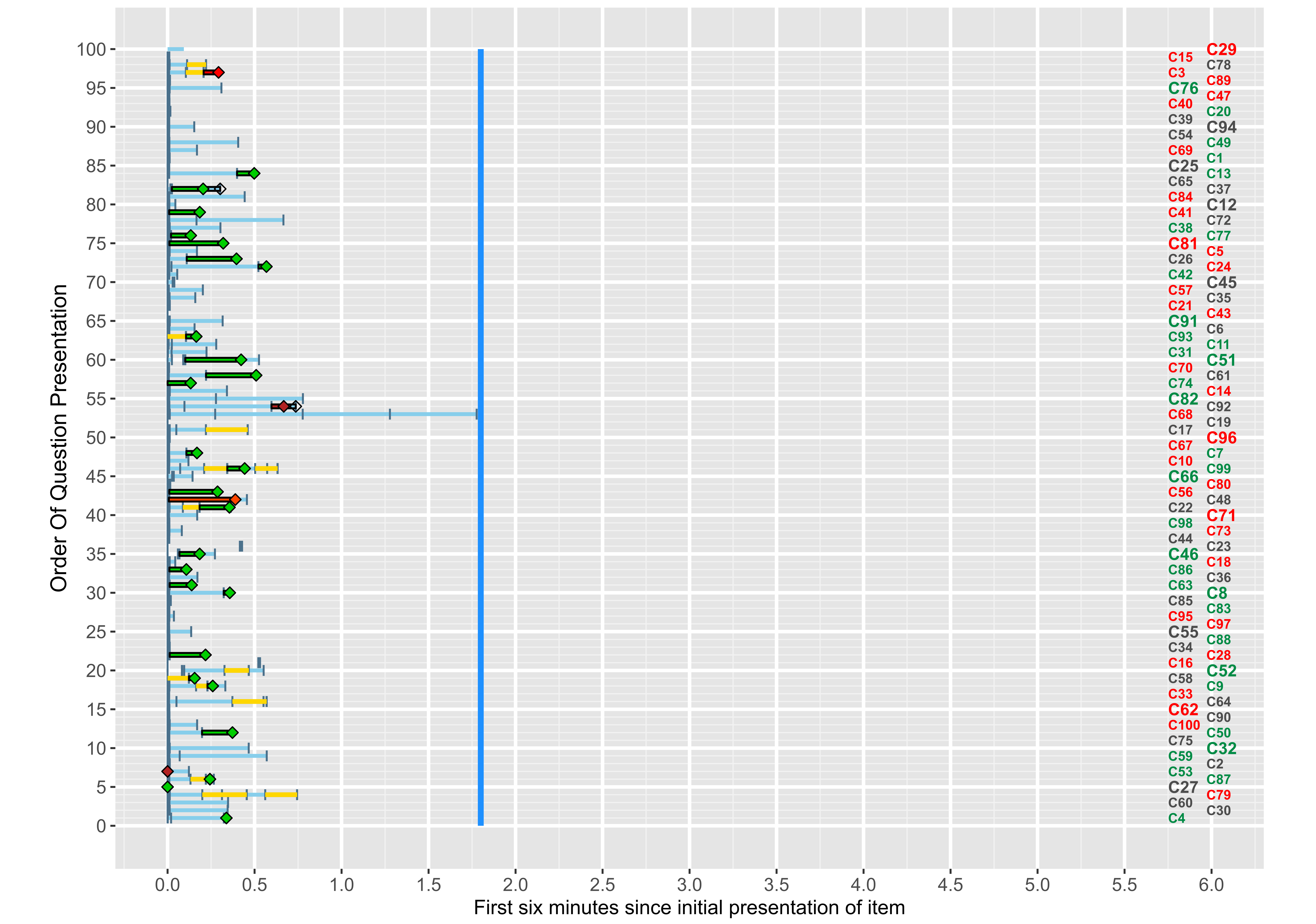

### Candidate_G_Full_ClickPlot.png

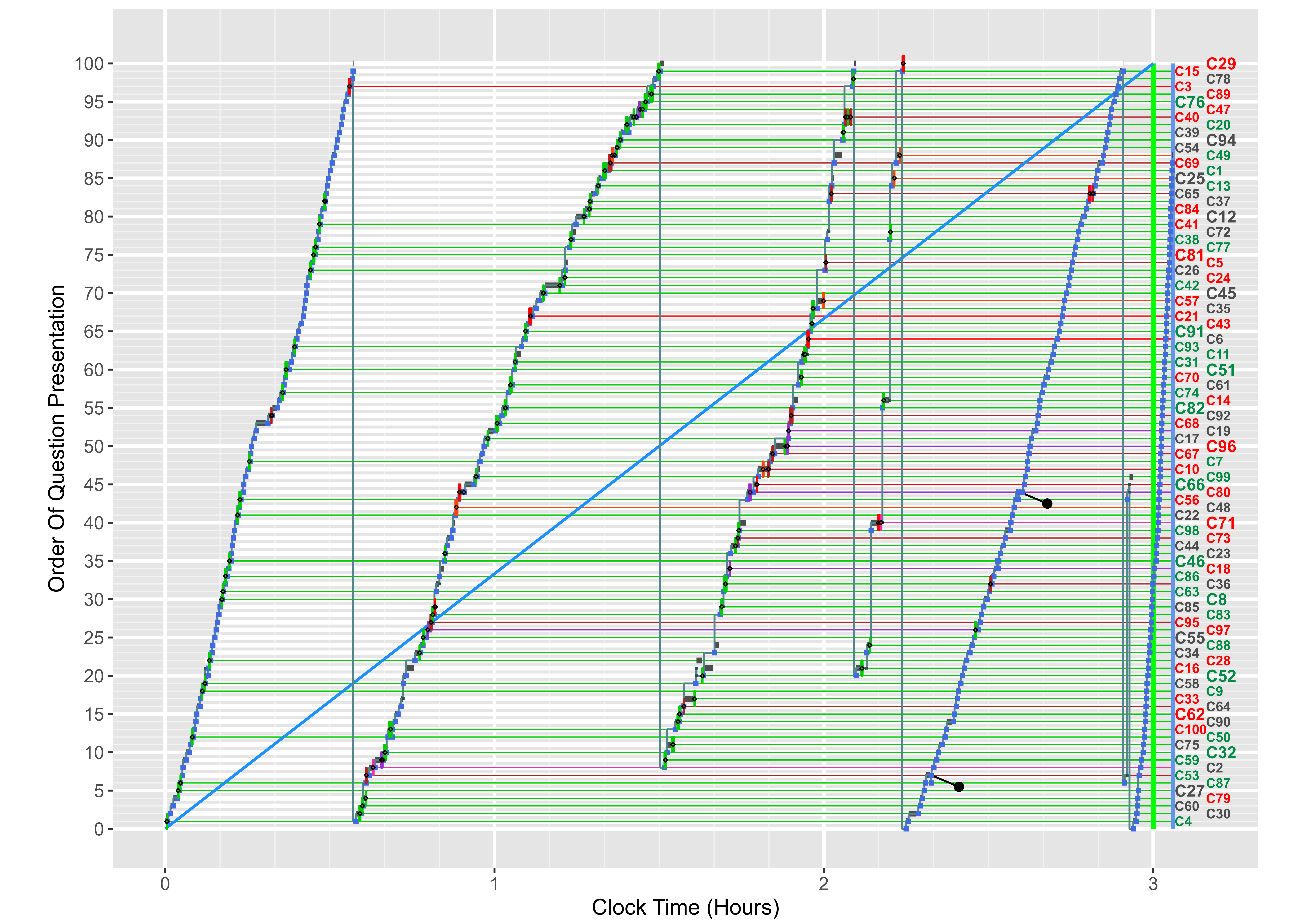

### Candidate_H_Early_ClickPlot.png

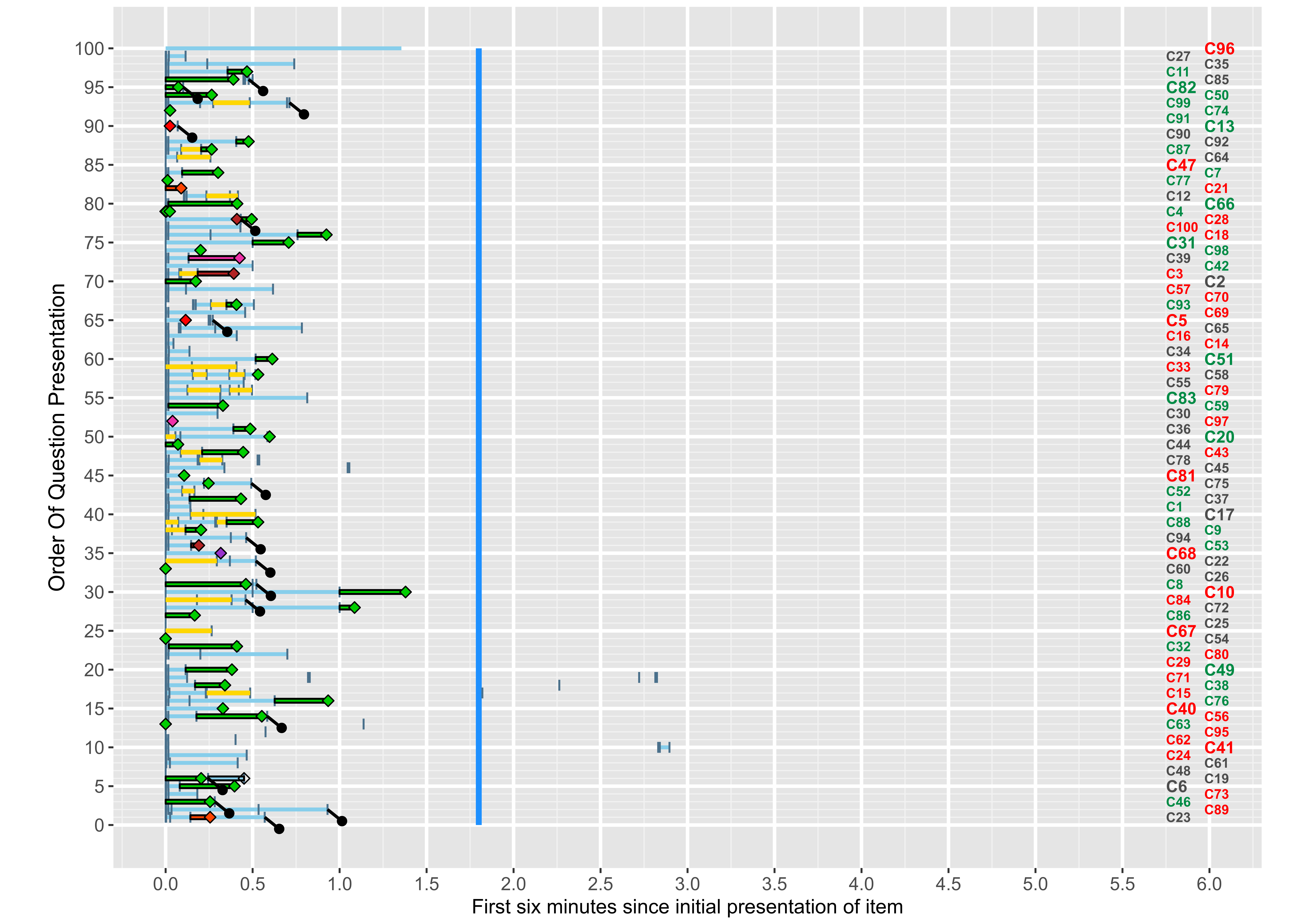

### Candidate_H_Full_ClickPlot.png

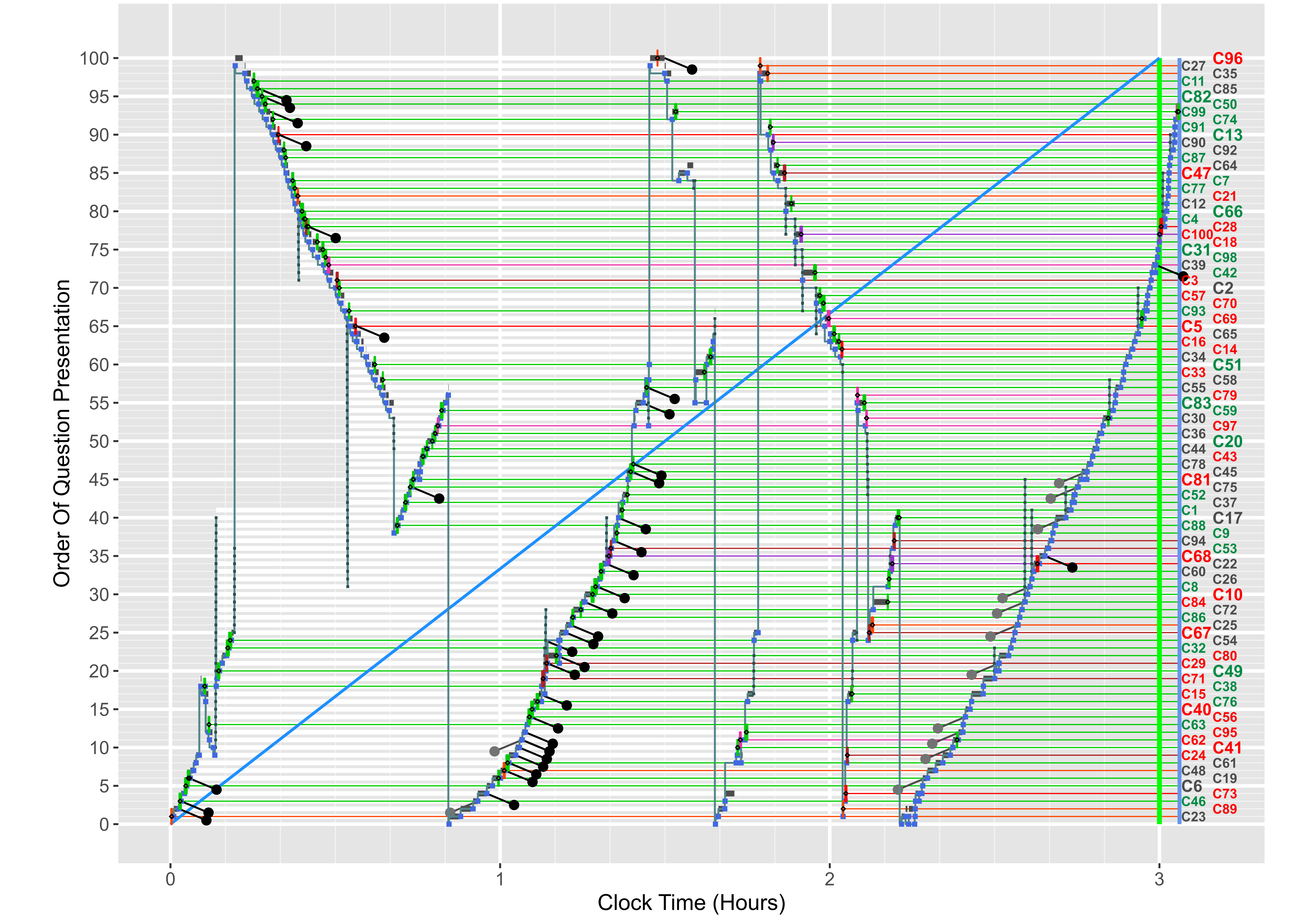

### Candidate_I_Early_ClickPlot.png

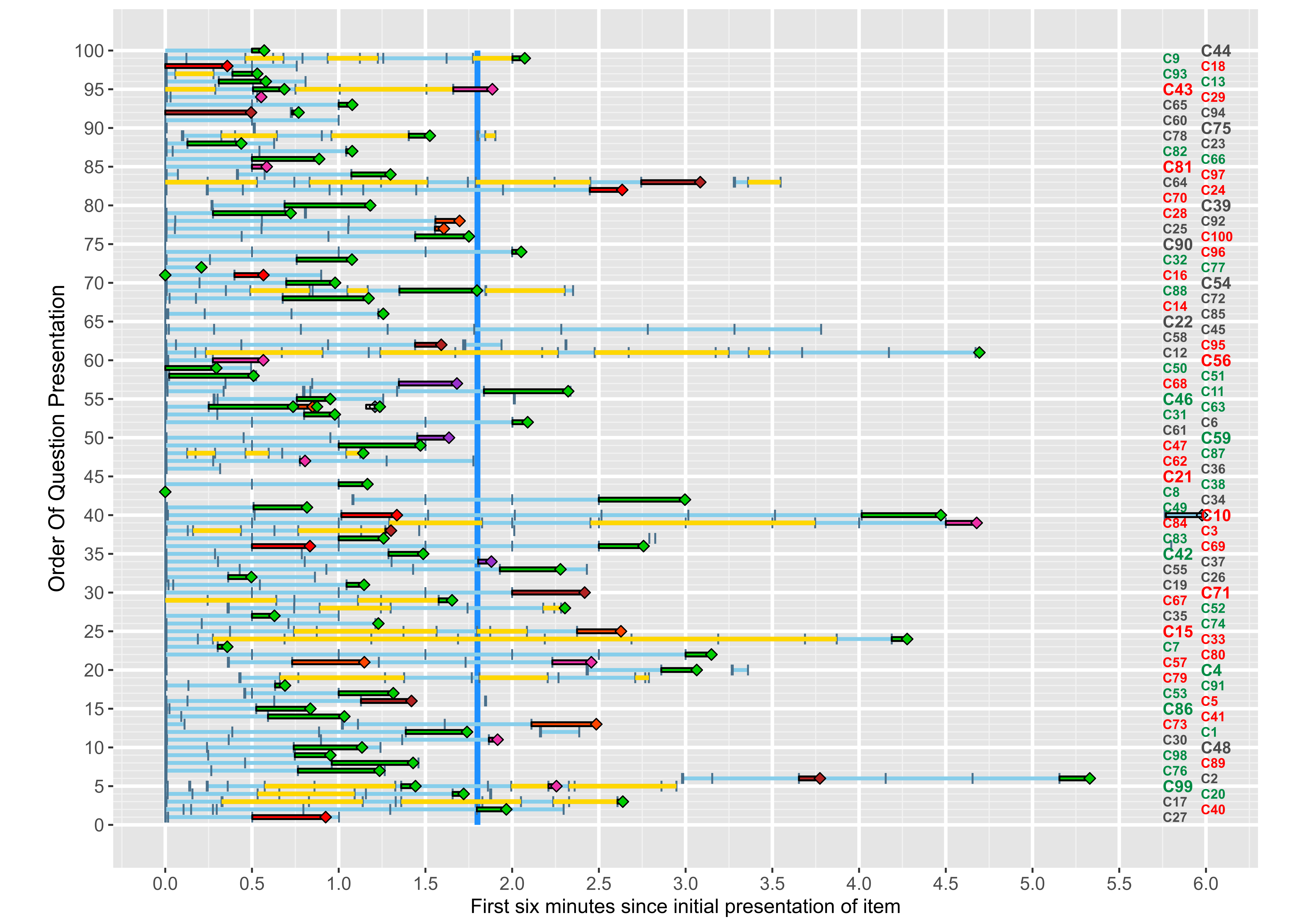

### Candidate_I_Full_ClickPlot.png

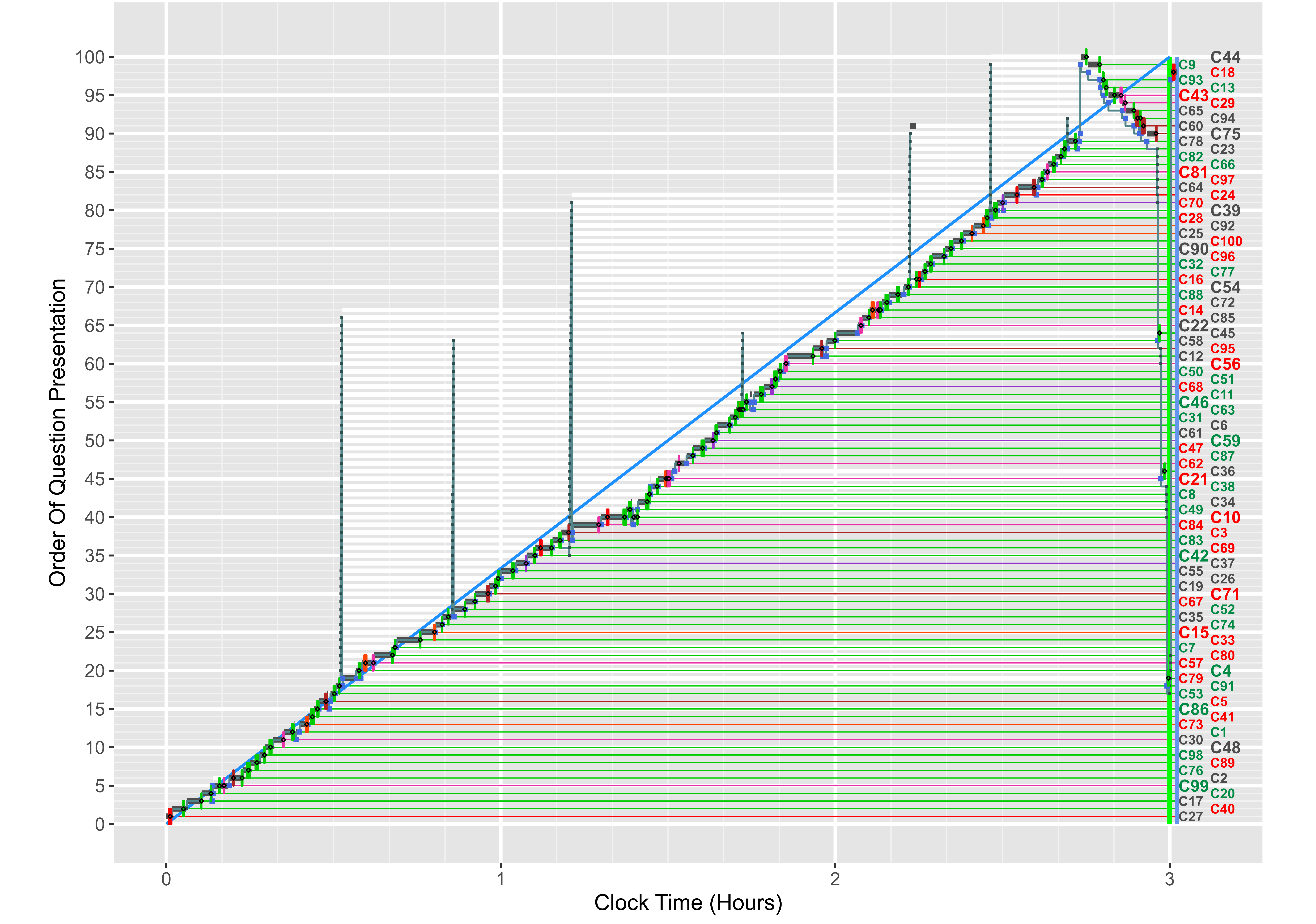

### Candidate_J_Early_ClickPlot.png

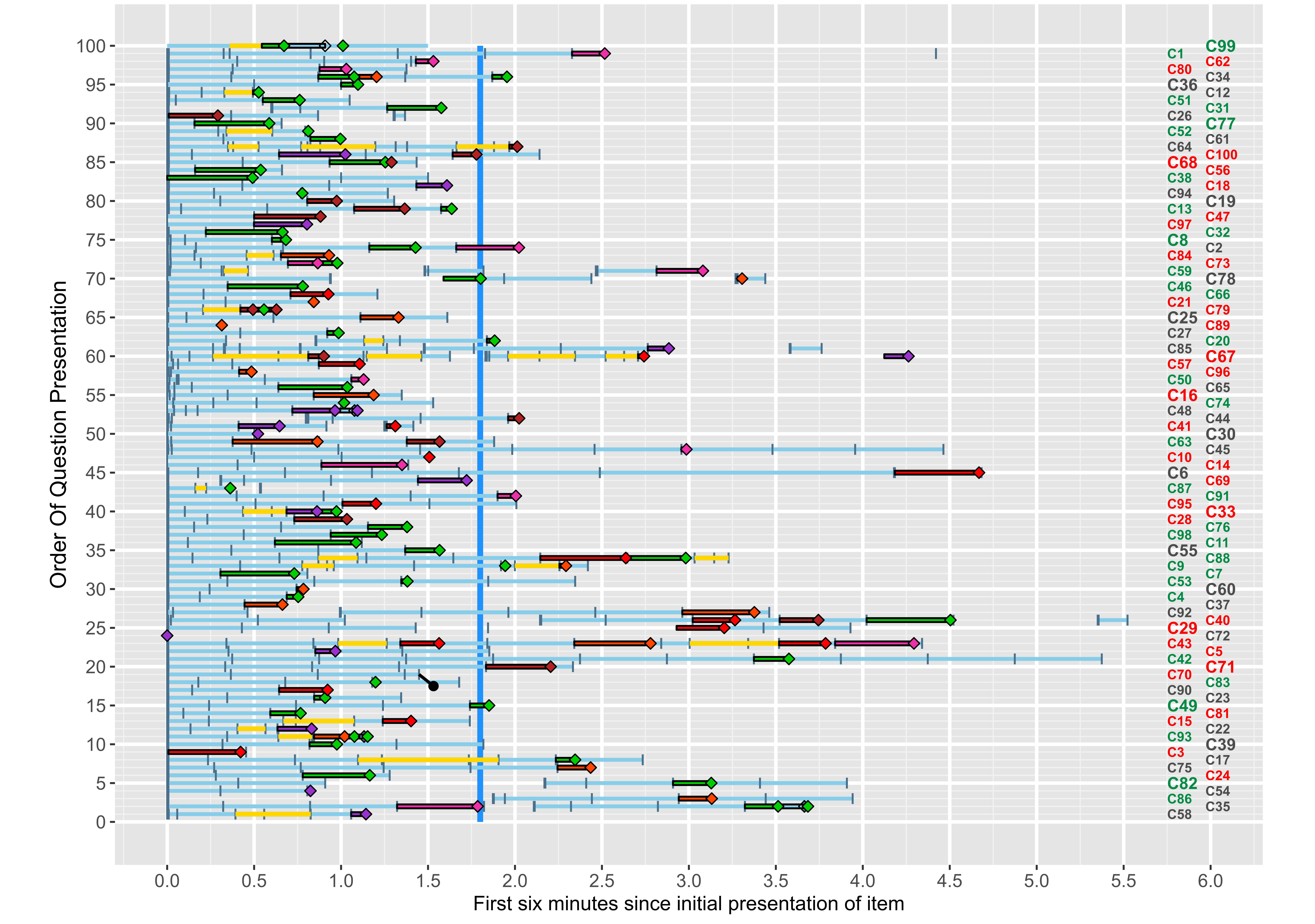

### Candidate_J_Full_ClickPlot.png

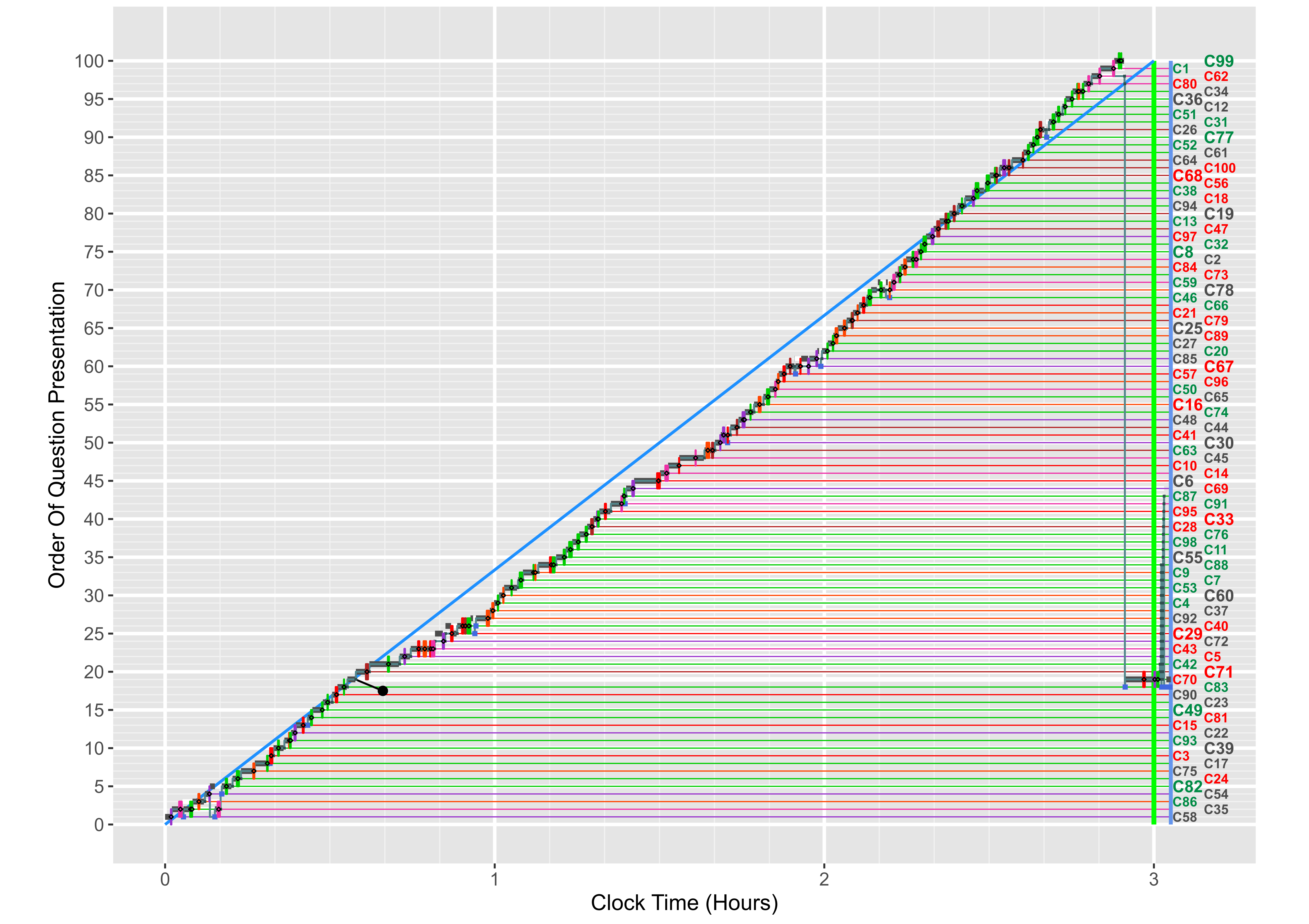

### Candidate_K_Early_ClickPlot.png

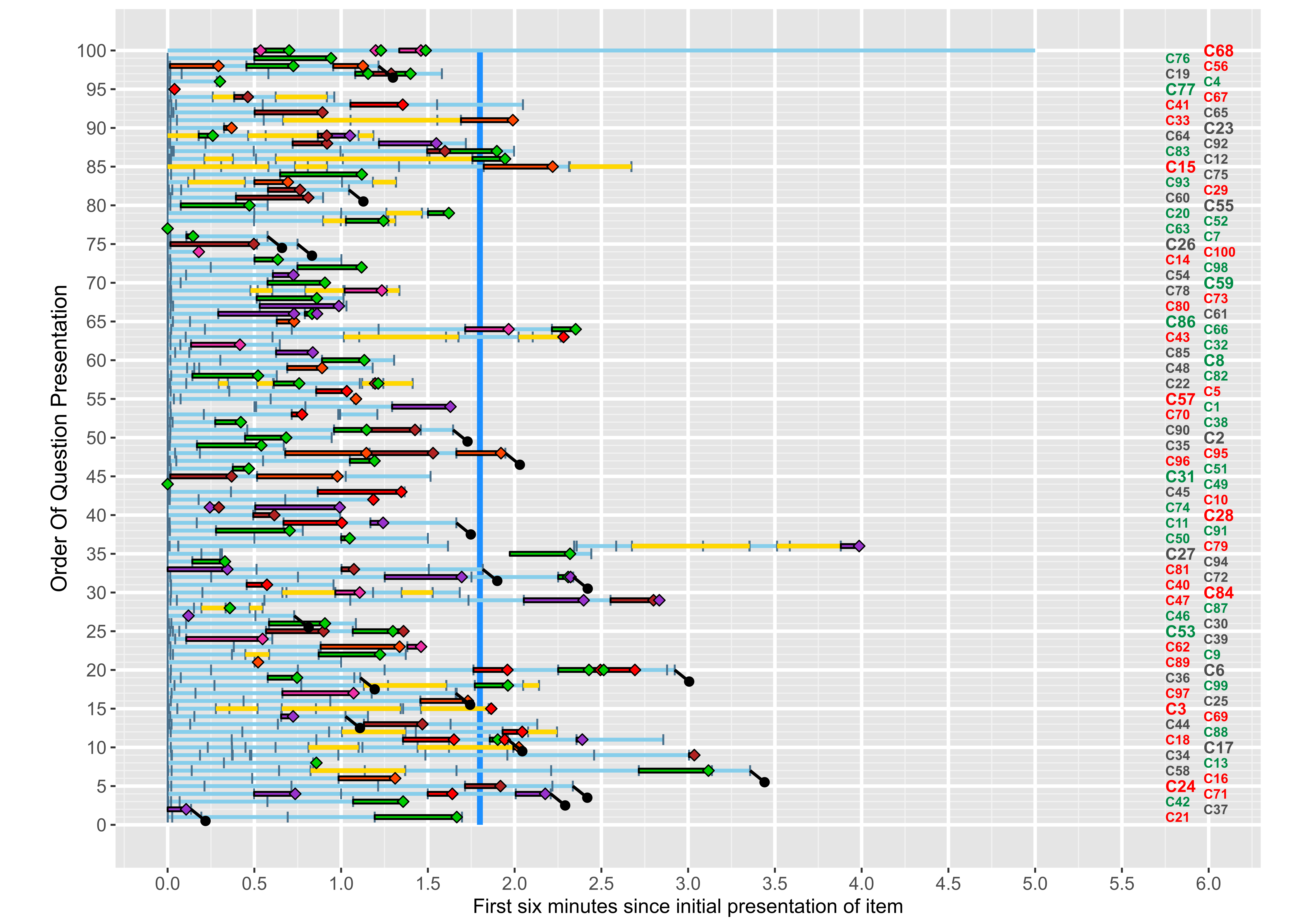

### Candidate_K_Full_ClickPlot.png

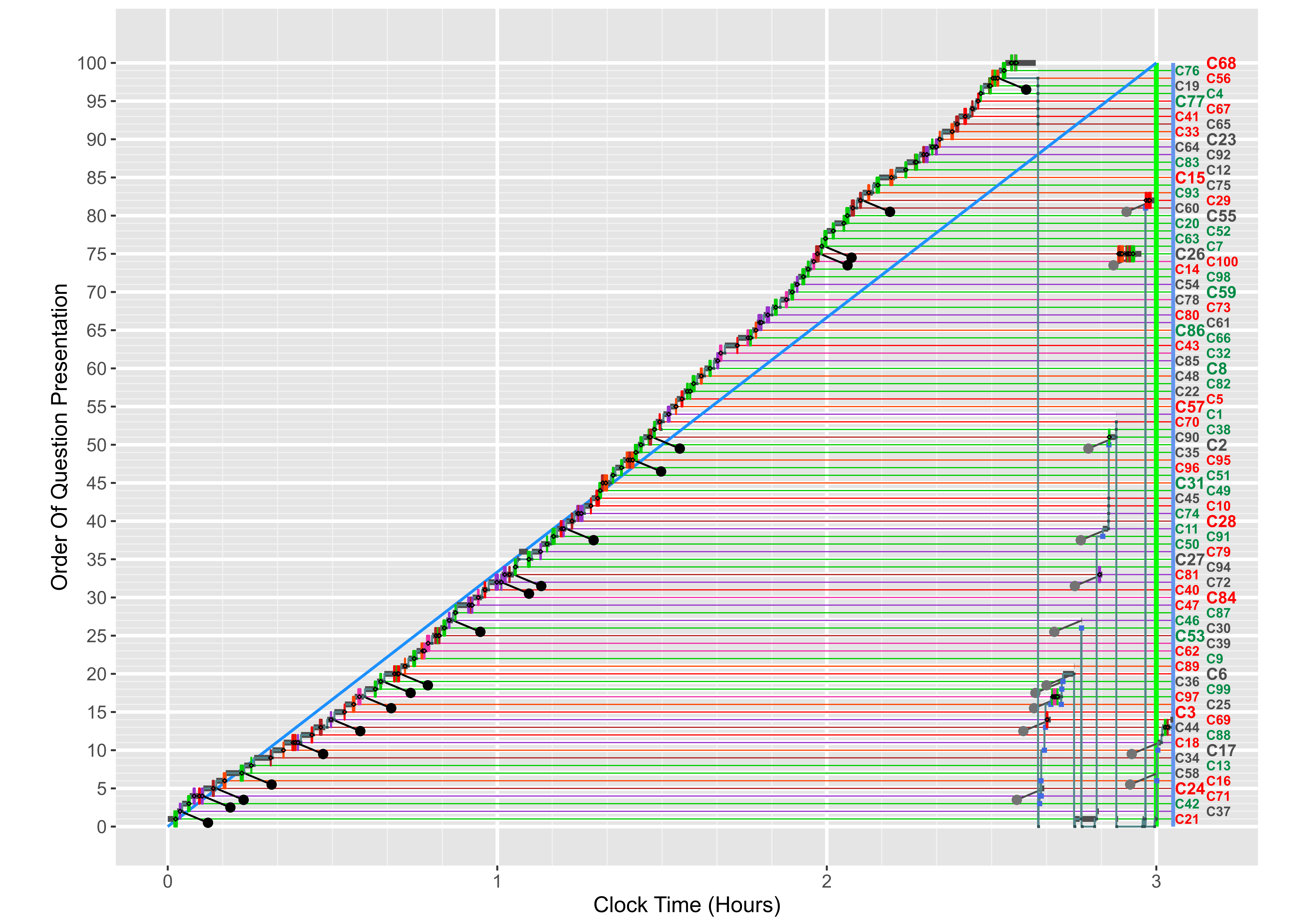
